# Symptom-led staging for primary progressive aphasia

**DOI:** 10.1101/2023.03.13.23286972

**Authors:** Chris JD Hardy, Cathleen Taylor-Rubin, Beatrice Taylor, Emma Harding, Aida Suarez Gonzalez, Jessica Jiang, Laura Thompson, Rachel Kingma, Anthipa Chokesuwattanaskul, Ffion Walker, Suzie Barker, Emilie Brotherhood, Claire Waddington, Olivia Wood, Nikki Zimmermann, Nuriye Kupeli, Keir XX Yong, Paul M Camic, Josh Stott, Charles R Marshall, Neil P. Oxtoby, Jonathan D Rohrer, Anna Volkmer, Sebastian J Crutch, Jason D Warren

**Author notes:** Corresponding author: Prof Jason D Warren, Dementia Research Centre, UCL Institute of Neurology, Queen Square, London WC1N 3BG, UK.

## Abstract

The primary progressive aphasias (PPA) present complex and diverse challenges of diagnosis, management and prognosis. A clinically-informed, syndromic staging system for PPA would take a substantial step toward meeting these challenges. This study addressed this need using detailed, multi-domain mixed-methods symptom surveys of people with lived experience in a large international PPA cohort.

We administered structured online surveys to caregivers of patients with a canonical PPA syndromic variant (nonfluent/agrammatic (nvPPA), semantic (svPPA) or logopenic (lvPPA)). In an ‘exploratory’ survey, a putative list and ordering of verbal communication and nonverbal functioning (nonverbal thinking, conduct and wellbeing, physical) symptoms was administered to 118 caregiver members of the UK national PPA Support Group. Based on feedback, we expanded the symptom list and created six provisional clinical stages for each PPA subtype. In a ‘consolidation’ survey, these stages were presented to 110 caregiver members of UK and Australian PPA Support Groups, and refined based on quantitative and qualitative feedback. Symptoms were retained if rated as ‘present’ by a majority (at least 50%) of respondents representing that PPA syndrome, and assigned to a consolidated stage based on majority consensus; the confidence of assignment was estimated for each symptom as the proportion of respondents in agreement with the final staging for that symptom. Qualitative responses were analysed using framework analysis.

For each PPA syndrome, six stages ranging from 1 (‘Very mild’) to 6 (‘Profound’) were identified; earliest stages were distinguished by syndromic hallmark symptoms of communication dysfunction, with increasing trans-syndromic convergence and dependency for basic activities of daily living at later stages. Spelling errors, hearing changes and nonverbal behavioural features were reported at early stages in all syndromes. As the illness evolved, swallowing and mobility problems were reported earlier in nfvPPA than other syndromes, while difficulty recognising familiar people and household items characterised svPPA and visuospatial symptoms were more prominent in lvPPA. Overall confidence of symptom staging was higher for svPPA than other syndromes. Across syndromes, functional milestones were identified as key deficits that predict the sequence of major daily life impacts and associated management needs. Qualitatively, we identified five major themes encompassing 15 subthemes capturing respondents’ experiences of PPA and suggestions for staging implementation.

This work introduces a prototypical, symptom-led staging scheme for canonical PPA syndromes: the PPA Progression Planning Aid (PPA^2^). Our findings have implications for diagnostic and care pathway guidelines, trial design and personalised prognosis and treatment for people living with these diseases.

## INTRODUCTION

The primary progressive aphasias (PPA) are a diverse group of language-led neurodegenerative dementias that collectively constitute a major cause of dementia in younger people and pose unique challenges for diagnosis and management^1–10^. Diagnosis of PPA is frequently delayed^11^, and even after a diagnosis is made there is no clear ‘roadmap’ for anticipating the development of deficits and disability and care planning for individual patients and families journeying through these diseases^4, 5^. A clinical staging system for PPA would provide such a roadmap. While the potential value of clinical staging in neurodegenerative diseases is widely acknowledged^12–16^, this enterprise faces several fundamental difficulties. In contrast to diseases such as cancer, assessing disease burden objectively in dementia is problematic; the key pathophysiological milestones of disease progression are often unknown, there is wide individual variation in phenotypic expression and the mapping of clinical signs and functional deficits onto the spread of underlying tissue pathology is incompletely specified. All these challenges are amplified in PPA, reflecting the intrinsic complexity of language functions, a comparative lack of reliable *in vivo* progression biomarkers and the marked heterogeneity of clinical phenotypes^2, 3,17–19^.

Three canonical syndromes of PPA are recognised, each with a distinct profile of clinico-anatomical features and disease associations. The nonfluent/agrammatic variant (nfvPPA) is led by impaired speech production linked to predominant left anterior peri-Sylvian cortical atrophy and frequent evolution of atypical parkinsonism due to an underlying primary neurodegenerative tauopathy; while the semantic variant (svPPA) is led by breakdown of vocabulary and semantic memory for nonverbal objects and concepts, linked to focal left anterior temporal atrophy and underlying TDP43 pathology; and in contrast, the logopenic variant (lvPPA) presents with anomia and reduced verbal short term memory, usually linked to left temporo-parietal degeneration as a manifestation of Alzheimer’s disease pathology^17, 20^. This formulation masks considerable individual variation in the profile of deficits and speed of disease progression, and excludes a substantial minority of cases not meeting criteria for a single canonical syndromic diagnosis, even early in the course of the illness^2, 3, 6, 21, 22^. On the other hand, there is a strong clinical impression that most patients with PPA do transition through differentiable stages of impairment and functional disability: a ‘stage’ here would be defined by a particular constellation of problems, developing as part of a sequence that is broadly similar among patients with a given syndromic diagnosis. Early on, there is often loss of facility with more formal or structured verbal exchanges and subtle changes in social behaviour, while late stage disease tends to be defined by motor and other physical impairments often accompanied by more profound behavioural changes. Clinical experience further suggests that the sequence of impairments (or stages) follows a trajectory that differs between canonical PPA syndromes, an impression substantiated by analyses of the published literature^11, 23^.

Existing staging instruments for PPA and other dementias rest primarily on concepts of disease severity and evolution formulated by clinicians, rather than the lived experience of patients and caregivers. Standard instruments for staging dementia such as the Mini-Mental State Examination^24^, the Global Deterioration Scale^13^ and the Mattis Dementia Rating Scale^25^ (developed for Alzheimer’s disease) do not assess language and communication functions adequately for this purpose in PPA. Clinical severity rating scales relevant to PPA have been developed, notably the Frontotemporal Dementia Rating Scale^14^, the Clinical Dementia Rating + National Alzheimer’s Disease Coordinating Center Frontotemporal Lobar Degeneration scale^26, 27^ and the Progressive Aphasia Severity Scale^28^. However, these scales have limitations in their coverage of the clinical spectrum of PPA. The Frontotemporal Dementia Rating Scale, for example, lacks granularity in the assessment of language and communication functions (and does not address one of the canonical PPA subtypes, lvPPA) while the Progressive Aphasia Severity Scale does not include non-language symptoms, which are often prominent in PPA and may dictate overall functional impact of the illness^6, 29–36^. Identification of the earliest symptoms of PPA is set to become increasingly urgent in the dawning era of disease modification, as early stage disease will present the greatest opportunities for effective intervention^37^.

Here we addressed these challenges by developing a ‘Primary Progressive Aphasia Progression Planning Aid’ (PPA^2^): a bespoke, symptom-led staging system that synthesises the ‘top-down’ expertise of researchers and clinicians and the ‘bottom-up’ perspectives of those with lived experience of PPA (Figure 1). We adopt the term ‘clinical stages’ throughout to signify that these are phenomenological, symptom-led descriptions of PPA progression, but envisage that the PPA^2^ will be of use to clinicians and people with lived experience of PPA. Data supporting development of the PPA^2^ were collected from a large, international cohort of English-speaking patients with PPA and their primary caregivers. The cohort represented all canonical syndromes and a wide spectrum of disease severity. Following approaches applied to other neurodegenerative diseases^38–40^, caregivers completed a structured survey on the development of symptoms in PPA, designed to tap their lived experience of the disease and sampling verbal, nonverbal cognitive, behavioural and somatic functional domains. Our objectives were firstly, to obtain a detailed, whole disease-course lived PPA experience-reported picture of the evolution of all major PPA syndromes, as reflected in patients’ daily life functioning; to collate survey data into accessible descriptions of different ‘stages’ of this evolution, covering all phases of the illness; to capture qualitatively caregivers’ impressions of illness impact and trajectory and the value of the staging exercise; and finally, to identify key clinical milestones of PPA evolution that are likely to signal major changes in daily life function and could direct decisions around support and care needs.

**Figure 1.**
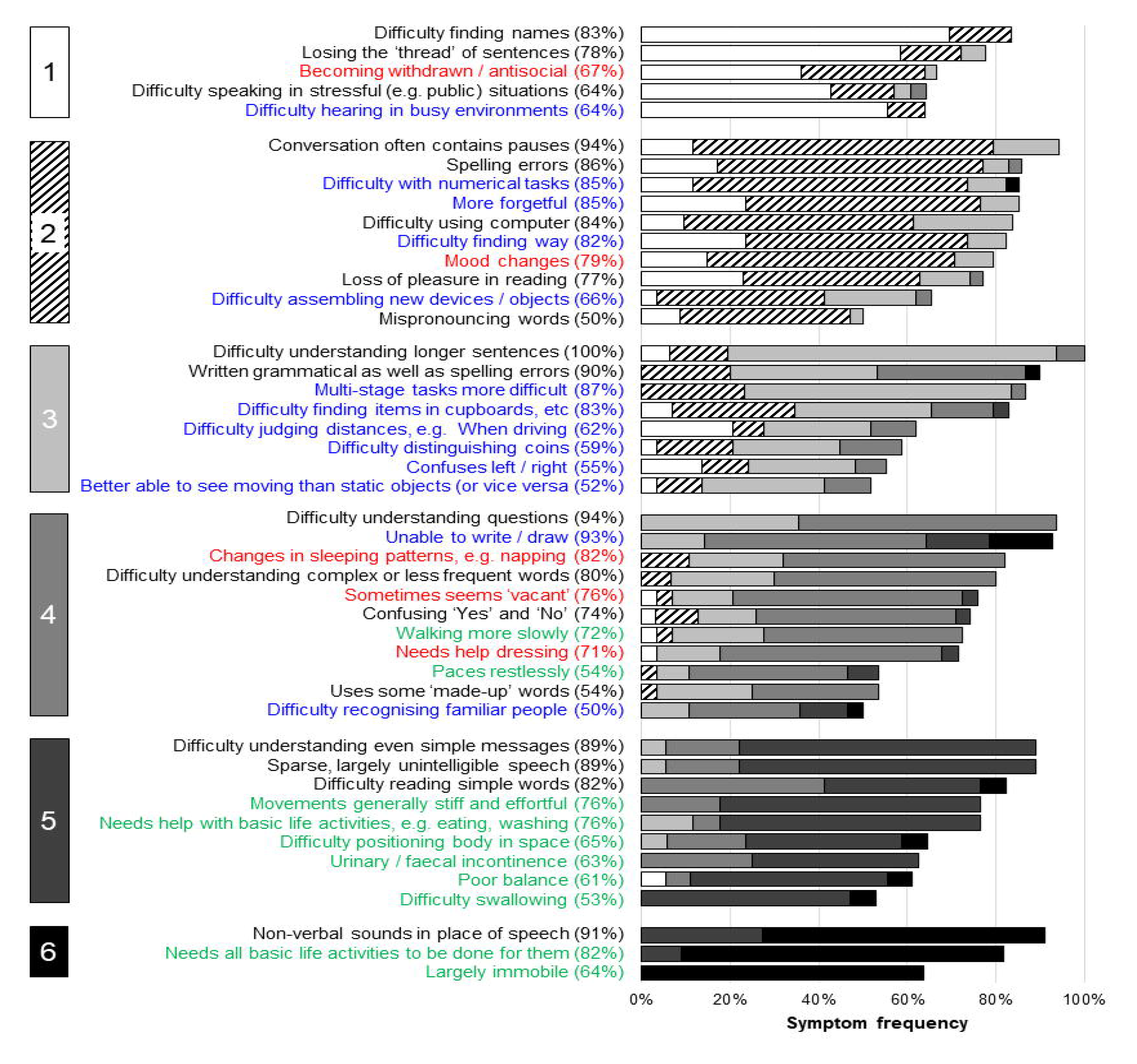
Development of the Primary Progressive Aphasia Progression Planning Aid (PPA^2^) The Figure illustrates the different sources of information incorporated in the Primary Progressive Aphasia Progression Planning Aid (PPA^2^). Data were synthesised from ‘top-down’ sources to characterise what was known from a clinician/researcher perspective (i.e. clinician-led interpretation of patient records, histories and neuropsychological test scores) and from ‘bottom-up’ sources to capture crucial information from those with lived experience of the conditions (i.e. patient/caregiver-derived symptoms and changes which were organised, prioritised and amended to reflect the lived experience of disease progression). The PPA^2^ puts forward two levels that clinicians and people with lived experience of PPA may find useful: the six stages give a granular overview of specific symptoms (see Figures 2-4); the three phases comprise broader markers containing key milestones with implications for management of these diseases (see Figure 5).

## METHODS

### Exploratory survey

An initial putative list and ordering of PPA symptoms was proposed by two of the authors (CJDH and JDW), based on clinical observations in the Dementia Research Centre PPA cohort and detailed review of case notes for patients with PPA attending the Cognitive Disorders Clinic at the National Hospital for Neurology and Neurosurgery. Symptoms covered domains of verbal communication (A) and nonverbal functioning (nonverbal thinking, B1; conduct and wellbeing, B2; physical, B3). Using an online survey hosted on the Opinio platform (www.objectplanet.com/opinio), qualitative feedback on the symptoms was gathered from 118 caregivers for people with a canonical syndrome of PPA who were registered with the United Kingdom national PPA Support Group (www.raredementiasupport.org). Data were collected between October 2018 and January 2019, and respondents comprised 41 caregivers of people with svPPA, 43 with nfvPPA and 34 with lvPPA; all had had longstanding personal contact with the patients whose illness they described. Based on this survey, we expanded the list of symptoms and created a provisional six-stage scheme for each PPA subtype. Stage labels were chosen to align with the Frontotemporal Dementia Rating Scale^14^: 1, ‘Very mild; 2, ‘Mild’; 3, ‘Moderate’; 4, ‘Severe’; 5, ‘Very severe’; 6, ‘Profound’. We added overarching descriptors pertaining to daily life and care implications for each stage, broadly based on the Reisberg Global Deterioration Scale^13^ (Table 1) as well as descriptors used previously in stages for another rare dementia, posterior cortical atrophy^41^; see Tables S1 to S3.

**Table 1.** Cross-syndromic descriptions of the PPA Progression Planning Aid (PPA^2^) stages

| PPA stage | Daily life impact | Care implications |
| --- | --- | --- |
| **1: Very mild** | This stage reflects the earliest clinical manifestations of PPA, but the person may not seek help, attributing problems to stress or ageing. Symptoms may be intermittent, difficult for the person to describe, or recognised only by those who know them well or in hindsight. | Definitive diagnosis of PPA may not be possible but clinical vigilance is required and specialist referral may be helpful |
| **2: Mild** | Communication and other less prominent problems with everyday activities are generally evident to others as well as to the person themselves (though sometimes insight may be lacking). | By this stage, a confident diagnosis of PPA by an experienced clinician should be possible |
| **3: Moderate** | The person may now require help managing certain aspects of day-to-day life and will generally have to stop working. Communication difficulties tend to frustrate important goals and social activities. | Engagement of supports and local services becomes essential |
| **4: Severe** | The person now requires support with many aspects of daily living and communication is increasingly difficult. They may no longer be able to live independently. | Care needs are complex and evolving, and safety concerns are paramount along with interventions to mitigate caregiver mental health issues and burnout |
| **5: Very severe** | Cognitive and behavioural changes are more global in nature, and many are common to all PPA syndromes. Meaningful communication is rarely possible. The person is likely to need help with daily personal care including toileting, and physical symptoms will have developed to the extent that mobility is significantly affected. | Significant complications and comorbidities may require management in their own right (e.g., the consequences of falls, immobility and/or incontinence) |
| **6: Profound** | Communication is now no longer possible. The person may lose their ability to respond to their environment and becomes largely immobile | Focus should now be placed on end-of-life care and bereavement support |

### Consolidation of the provisional stages

The provisional stages for each PPA syndrome were next entered into another online, ‘consolidation’ survey, designed to allow us to refine the provisional staging scheme. This second survey was refined for comprehensibility and presentation based on feedback from the exploratory survey and published guidelines for online research survey design^42^, and was again hosted on the Opinio platform (www.objectplanet.com/opinio). This voluntary survey was distributed via email to caregivers comprising members of the UK PPA Support Group and PPA Support Groups in Melbourne and Sydney, Australia. Both current and bereaved caregivers were surveyed, to allow us to include information about late stage disease, and data were collected between February 2020 and April 2020 for UK Support Group respondents and between January 2021 and May 2021 for Australian Support Group respondents. All caregivers had again had longstanding personal contact with the patients whose illness they described.

At the top of the survey, caregiver respondents first identified the major syndromic diagnosis for which they were filling the survey and provided information about their relationship to the patient, and the patient’s age currently, at symptom onset, when first assessed medically and when diagnosed. The symptom labels presented to respondents in the consolidation survey are given in full in Supplementary Material online (Table S1 to S3). Customised symptom lists were presented under each stage according to the syndromic PPA diagnosis with which the respondent self-identified at the top of the survey: this was to ensure respondent caregivers were able to focus on symptoms most relevant to ‘their’ syndrome, while keeping their task manageable. A given PPA stage will be defined by a particular conjunction of symptoms, however, there was no prior ‘ground truth’ to determine the correct conjunction for each stage. For each symptom, survey respondents were therefore asked to indicate whether, based on proximity to other symptoms and the overarching stage description (Table 1), the symptom began at the stage to which it was provisionally assigned, if it began at an earlier or a later stage (and which one), or if it was absent altogether (i.e., the respondent did not recognise that symptom as ever having been experienced over the course of the patient’s illness to date). We assumed that respondents for patients who were earlier in the course of their illness would not recognise most symptoms assigned provisionally to later PPA stages; moreover, we wished to avoid causing distress by confronting respondents with symptoms they might not have anticipated. Respondents were therefore able to discontinue this first section of the survey at any point. The point at which the respondent discontinued this section of the survey was taken to indicate that patient’s current PPA stage. Participants were able to review and edit their responses at any point via a ‘Back’ button.

In the next section of the survey, respondents were presented with a representative list of symptoms present i) in other forms of PPA (sampling each of the domains A, B1, B2 and B3), and ii) (principally as an internal ‘control’, to asses response bias) in a staging system for a clinically distinct, ‘visual’ dementia (posterior cortical atrophy, PCA)^41^: for each of these symptoms, caregivers were again asked to indicate whether the symptom was present, and if so, to which stage it should be assigned. They were additionally given the opportunity to make additional comments about symptoms not covered elsewhere in the survey, and their impressions of the staging system in its current form, for the purpose of qualitative analysis.

### Validation of diagnosis

To allow us to estimate the overall validity of syndromic diagnoses as listed by caregivers in the consolidation survey, survey respondents recruited from the UK PPA Support Group were given the option of including their name when they completed the survey. When this was volunteered, we were able to cross-check whether that caregiver-patient dyad had previously participated in the PPA research programme at Queen Square; and if they had, to check that the diagnosis listed in the survey for that person living with PPA was corroborated by detailed neuropsychological and neuroimaging data held on the research database.

### Analysis of clinical and demographic data

Clinical and demographic data were analysed using JASP version 0.16.2^43^. Groups (i.e. variants) were compared using ANOVAs for continuous variables and Fisher’s exact tests for categorical variables.

### Quantitative analysis of survey responses

For each symptom in the consolidation survey, we calculated the percentage of respondents who had declared that symptom to be ‘present’, regardless of PPA subtype or stage; a symptom was retained only if a majority (at least 50%) of caregivers who provided a response to a given symptom reported it was present at some stage. In addition, we calculated the percentage of respondents who considered each symptom had been assigned to the correct stage. If a majority of respondents considered a symptom should be reassigned to an earlier or later stage, it was reassigned accordingly. Where a symptom was assigned to more than one stage (e.g. the majority was tied across two stages), it was retained only at the earlier stage for which it first achieved criterion (since in general, the earliest appearance of a symptom is most informative for signalling disease progression and/or planning care needs). We assessed the ‘confidence’ of stage assignment for each symptom as the proportion of respondents for that symptom in agreement with the final stage to which the symptom was assigned.

### Qualitative analysis of survey responses

Caregiver comments on the exploratory and consolidation surveys were analysed qualitatively using framework analysis^44^. A tentative framework was proposed by one of the authors (CJDH) following familiarisation with the whole dataset. This initial coding framework was then applied to a sub-sample of 20% of the dataset, which was then reviewed by another author (EH); discrepancies or differing interpretations were reviewed and discussed. Based on this consensus, a thematic framework was developed and applied to the full survey dataset.

### Ethical approval

Data for this study from UK PPA Support Group members were collected under the Rare Dementia Support (RDS) Impact Study protocol, which has been published separately^45^. Ethical approval was granted by the University College London Research Ethics Committee (8545/004: Rare Dementia Support (RDS) Impact study). Additional local site approval for Support Group members in Sydney was granted by the South Eastern Sydney Local Health District HREC (2020/ETH02530). All survey respondents gave informed consent, in accordance with Declaration of Helsinki guidelines. Survey data were downloaded from the online platform and stored within the UCL Data Safe Haven to protect against unauthorised access.

### Data availability

The data that support the findings of this study are available on request from the corresponding author. The data are not publicly available as they include information that could compromise the privacy of the research participants.

## RESULTS

The final stages for each PPA syndrome are presented in Figures 2 to 4, with overarching descriptors for each stage in Table 1. The raw data supporting the stage assignments are presented in Supplementary Tables S1 to S3 online. Themes, subthemes and illustrative caregiver comments from the qualitative framework analysis are presented in Tables 3 (reduced version) and S4 (full version).

**Figure 2.**
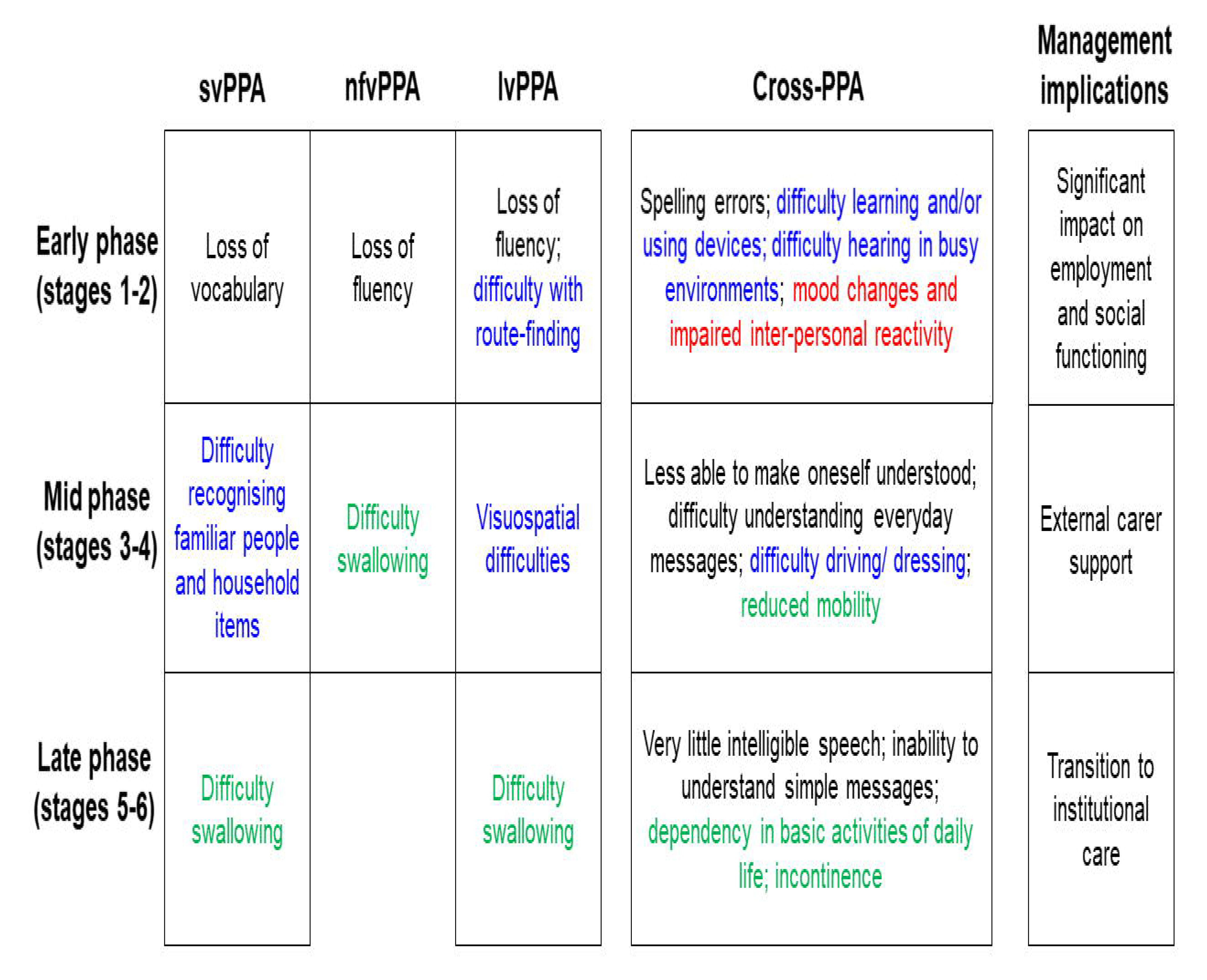
Symptom frequencies and confidence in symptom placement by stage for semantic variant primary progressive aphasia. The Figure shows all symptoms included in the svPPA Progression Planning Aid (see Supplementary Table S1). Boxes on the left-hand side denote stages (1 = very mild svPPA; 2 = mild svPPA; 3 = moderate svPPA; 4 = severe svPPA; 5 = very severe svPPA; 6 = profound svPPA). Written symptom labels are colour-coded based on domains of verbal communication (A = black) and nonverbal functioning (B1 = nonverbal thinking, blue; B2 = conduct and wellbeing, red); B3 = physical, green). Horizontal bars indicate the ‘confidence’ of symptom staging, calculated as the percentage of people responding to a given symptom who endorsed placement of that symptom in its final stage (i.e. the highest agreement achieved for placement of that symptom). Symptoms have been ordered within stages in descending order of overall frequency.

**Figure 3.**
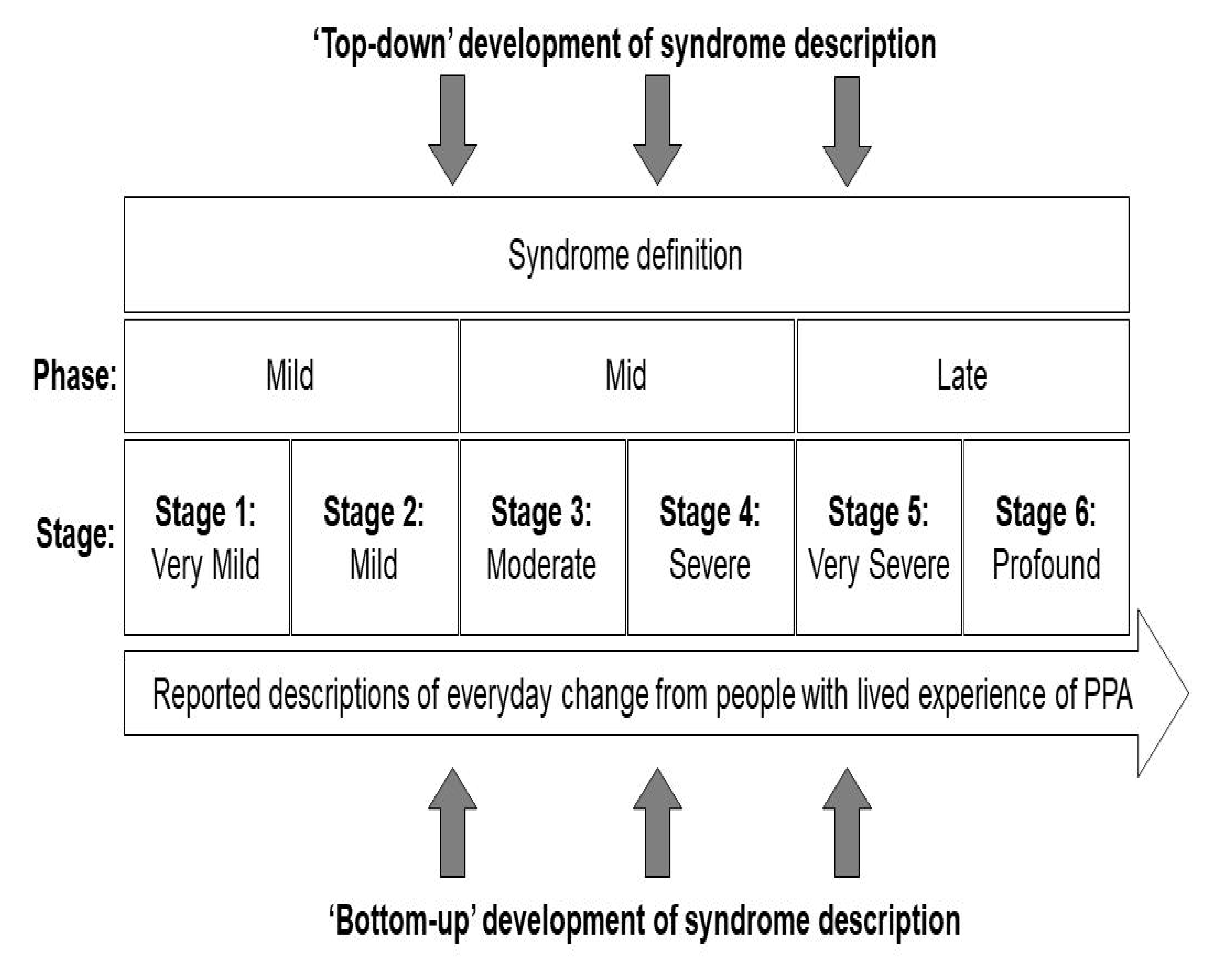
Symptom frequencies and confidence in symptom placement by stage for nonfluent-agrammatic variant primary progressive aphasia. The Figure shows all symptoms included in the nfvPPA Progression Planning Aid (see Supplementary Table S2. Boxes on the left-hand side denote stages (1 = very mild nfvPPA; 2 = mild nfvPPA; 3 = moderate nfvPPA; 4 = severe nfvPPA; 5 = very severe nfvPPA; 6 = profound nfvPPA). Written symptom labels are colour-coded based on domains of verbal communication (A = black) and nonverbal functioning (B1 = nonverbal thinking, blue; B2 = conduct and wellbeing, red); B3 = physical, green). Horizontal bars indicate the ‘confidence’ of symptom staging, calculated as the percentage of people responding to a given symptom who endorsed placement of that symptom in its final stage (i.e. the highest agreement achieved for placement of that symptom). Symptoms have been ordered within stages in descending order of overall frequency.

**Figure 4.**
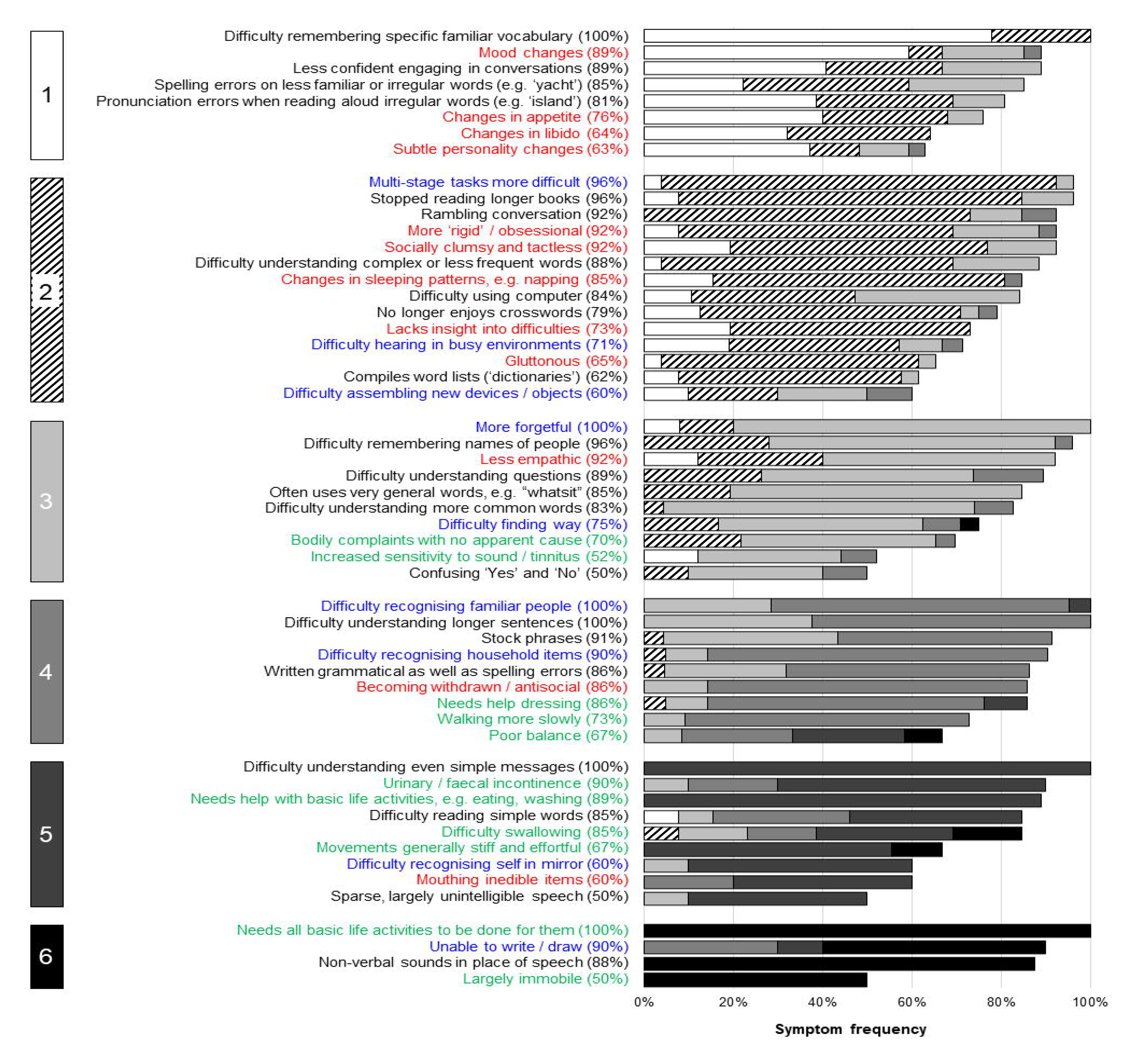
Symptom frequencies and confidence in symptom placement by stage for logopenic variant primary progressive aphasia. The Figure shows all symptoms included in the lvPPA Progression Planning Aid (see Supplementary Table S3. Boxes on the left-hand side denote stages (1 = very mild lvPPA; 2 = mild lvPPA; 3 = moderate lvPPA; 4 = severe lvPPA; 5 = very severe lvPPA; 6 = profound lvPPA). Written symptom labels are colour-coded based on domains of verbal communication (A = black) and nonverbal functioning (B1 = nonverbal thinking, blue; B2 = conduct and wellbeing, red); B3 = physical, green). Horizontal bars indicate the ‘confidence’ of symptom staging, calculated as the percentage of people responding to a given symptom who endorsed placement of that symptom in its final stage (i.e. the highest agreement achieved for placement of that symptom). Symptoms have been ordered within stages in descending order of overall frequency.

### Demographic and clinical characteristics

Demographic and clinical characteristics of patients whose data were provided for the caregiver consolidation survey are summarised in Table 2. The survey was accessed two hundred and six times, but data were removed for the following (not mutually exclusive) reasons: no response provided beyond agreeing to take part, n = 84; manually removed because data provided were atypical (e.g. respondent indicated that the person they were answering the survey on behalf of did not have a canonical PPA variant), n = 3; data directly duplicated a previous response, n = 3; the respondent was a professional caregiver, n = 1; and/or the person completing the survey had a diagnosis of PPA themselves, n = 7. After exclusions, responses were received from 110 primary caregivers: 27 acting for people with svPPA, 46 for people with nfvPPA and 37 for people with lvPPA, giving an overall participation rate of 53.9%.

**Table 2.**
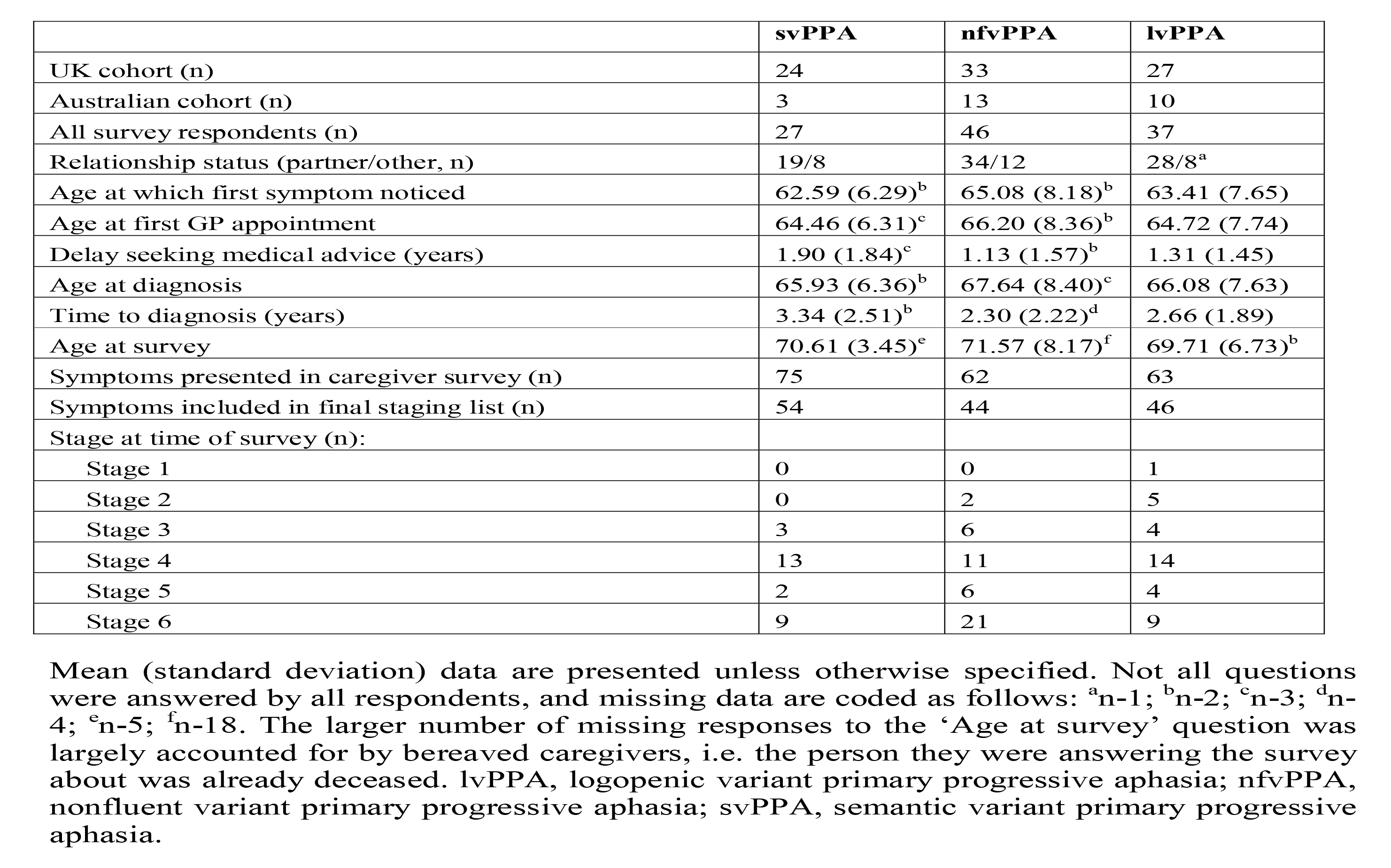
Breakdown of general respondent characteristics in the caregiver consolidation survey

|  | **svPPA** | **nfvPPA** | **lvPPA** |
| --- | --- | --- | --- |
| UK cohort (n) | 24 | 33 | 27 |
| Australian cohort (n) | 3 | 13 | 10 |
| All survey respondents (n) | 27 | 46 | 37 |
| Relationship status (partner/other, n) | 19/8 | 34/12 | 28/8<sup>a</sup> |
| Age at which first symptom noticed | 62.59 (6.29)<sup>b</sup> | 65.08 (8.18)<sup>b</sup> | 63.41 (7.65) |
| Age at first GP appointment | 64.46 (6.31)<sup>c</sup> | 66.20 (8.36)<sup>b</sup> | 64.72 (7.74) |
| Delay seeking medical advice (years) | 1.90 (1.84)<sup>c</sup> | 1.13 (1.57)<sup>b</sup> | 1.31 (1.45) |
| Age at diagnosis | 65.93 (6.36)<sup>b</sup> | 67.64 (8.40)<sup>c</sup> | 66.08 (7.63) |
| Time to diagnosis (years) | 3.34 (2.51)<sup>b</sup> | 2.30 (2.22)<sup>d</sup> | 2.66 (1.89) |
| Age at survey | 70.61 (3.45)<sup>e</sup> | 71.57 (8.17)<sup>f</sup> | 69.71 (6.73)<sup>b</sup> |
| Symptoms presented in caregiver survey (n) | 75 | 62 | 63 |
| Symptoms included in final staging list (n) | 54 | 44 | 46 |
| Stage at time of survey (n): |  |  |  |
| Stage 1 | 0 | 0 | 1 |
| Stage 2 | 0 | 2 | 5 |
| Stage 3 | 3 | 6 | 4 |
| Stage 4 | 13 | 11 | 14 |
| Stage 5 | 2 | 6 | 4 |
| Stage 6 | 9 | 21 | 9 |

Mean (standard deviation) data are presented unless otherwise specified. Not all questions were answered by all respondents, and missing data are coded as follows: <sup>a</sup>n-1; <sup>b</sup>n-2; <sup>c</sup>n-3; <sup>d</sup>n-4; <sup>e</sup>n-5; <sup>f</sup>n-18. The larger number of missing responses to the ‘Age at survey’ question was largely accounted for by bereaved caregivers, i.e. the person they were answering the survey about was already deceased. lvPPA, logopenic variant primary progressive aphasia; nfvPPA, nonfluent variant primary progressive aphasia; svPPA, semantic variant primary progressive aphasia.

Diagnostic groups for surveyed respondents did not differ significantly in age at symptom onset (F(2,103)=0.98, p=0.337), age at first visit to GP (F(2,102)=0.55, p=0.580), delay seeking medical advice (calculated as age at first visit to GP minus age at symptom onset; F(2,102)=1.80, p=0.170), age at diagnosis (F(2,102)=0.57, p=0.569), time to diagnosis (calculated as age at diagnosis minus age at symptom onset; F(2,100)=1.58, p=0.210), age when survey completed (F(2,80)=0.60, p=0.552), relationship status (Fisher’s exact p=0.785) or stage at the time of the survey (Fisher’s exact p=0.218). In all syndromes, the illness tended to declare itself from the early to mid-seventh decade, and with substantial individual variation (Table 2). For 32 self-identified respondent caregivers of people with PPA also participating in the Queen Square research programme, review of available neuropsychological and neuroimaging data in the research database confirmed the syndromic diagnosis listed in the survey in all cases. The average delay from symptom onset to diagnosis was well over two years across PPA syndromes, albeit with a wide range in each syndrome.

### Quantitative analysis of survey responses

More symptoms were endorsed overall at the frequency criterion (≥50%) by caregiver respondents for svPPA (54 symptoms) than for nfvPPA (44 symptoms) or lvPPA (46 symptoms). However, the six-stage scheme (ranging from stage 1, ‘Very mild’ to stage 6, ‘Profound’; Table 1) was endorsed in each PPA syndrome, and the overall profile of symptom development across domains was broadly similar for different syndromes. In all syndromes, symptoms relating to communication and non-verbal conduct and wellbeing were present at stage 1 and symptoms relating to nonverbal thinking by stage 2. Earliest symptoms included erosion of specific vocabulary in svPPA, difficulty conversing in stressful situations in both nfvPPA and lvPPA, and binary (e.g., ‘Yes’ / ‘No’) reversals in nfvPPA; spelling errors were an early feature in all syndromes. Difficulty hearing in busy environments also developed early in all syndromes and was among the first symptoms endorsed for lvPPA. Changes in inter-personal behaviour – altered libido in svPPA and nfvPPA and social withdrawal in lvPPA – were also among the earliest nonverbal features across syndromes, while loss of insight was endorsed for both svPPA and nfvPPA. Problems with episodic memory, route finding, praxis and task sequencing developed in all syndromes by stage 3, while visuospatial dysfunction was relatively more prominent in lvPPA; of 15 additional ‘control’ symptoms relevant to PCA presented in the survey, six (reflecting nonverbal parietal lobe functions, i.e. relating to praxis and visuoperceptual awareness) were endorsed by caregivers for inclusion in the stages for a PPA syndrome (most frequently in lvPPA; details in Tables S1 to S3). Physical symptoms began in stage 3 for nfvPPA and svPPA and stage 4 for lvPPA: the nature of these first physical symptoms varied between syndromes, patients with nfvPPA developing difficulties moving and swallowing and patients with svPPA unexplained somatic complaints, hyperacusis and tinnitus. The syndromic specificity of symptoms diminished substantially over the course of the illness (examining all symptoms listed in stages 1 and 2 across syndromes, 48% were unique to one variant, whereas this was the case for just 16% of symptoms listed in stages 5 and 6). End-stage PPA in all syndromes was characterised by vocal production limited to sparse, nonverbal sounds, inability to understand others, immobility and complete dependency for basic activities of daily life.

There was a wide range of ‘confidence’ in symptom placement, across stages and syndromes (Figures 2 to 4; Tables S1 to S3). For svPPA, overall respondent agreement with symptom placement increased over stages (mean 55% in stage 1, 89% in stage 6). In contrast, for nfvPPA, overall respondent agreement with staging remained moderate across stages (mean 53% in stage 1, 51% in stage 6); while for lvPPA, respondent agreement with symptom staging was good for early and late stages but reduced at intermediate stages (mean 73% in stage 1, 53% in stage 3, 86% in stage 6).

### Clinical milestones of PPA evolution

The grouping of symptoms under each stage suggested broad functional milestones in the evolution of PPA syndromes, raising management implications relevant to overarching disease ‘phases’ which we designate ‘early’ (stages 1 and 2), ‘mid’ (stages 3 and 4), and ‘late’ (stages 5 and 6) (Figure 5). Certain milestone symptoms at a given phase of the illness were associated with particular syndromes, while others occurred across all syndromes. In early phase disease, syndrome-specific milestone symptoms comprising loss of vocabulary in svPPA, loss of fluency in nfvPPA and lvPPA and route finding difficulty in lvPPA, coupled with cross-syndromic hearing, mood and socio-emotional changes, are predicted to impact occupational and/or social functioning. In mid phase disease, syndrome-specific milestone **symptoms** comprising difficulty recognising people and household items in svPPA, difficulty swallowing in nfvPPA and visuospatial difficulties in lvPPA, coupled with cross-syndromic difficulties understanding simple messages, driving, dressing, and/or mobilising, would predict an increasing need for external care supports. In late-phase disease, the development of dysphagia in svPPA and lvPPA coupled with cross-syndromic loss of communication function, dependency in basic activities of daily life and incontinence, might raise the prospect of a transition to residential care.

**Figure 5.**
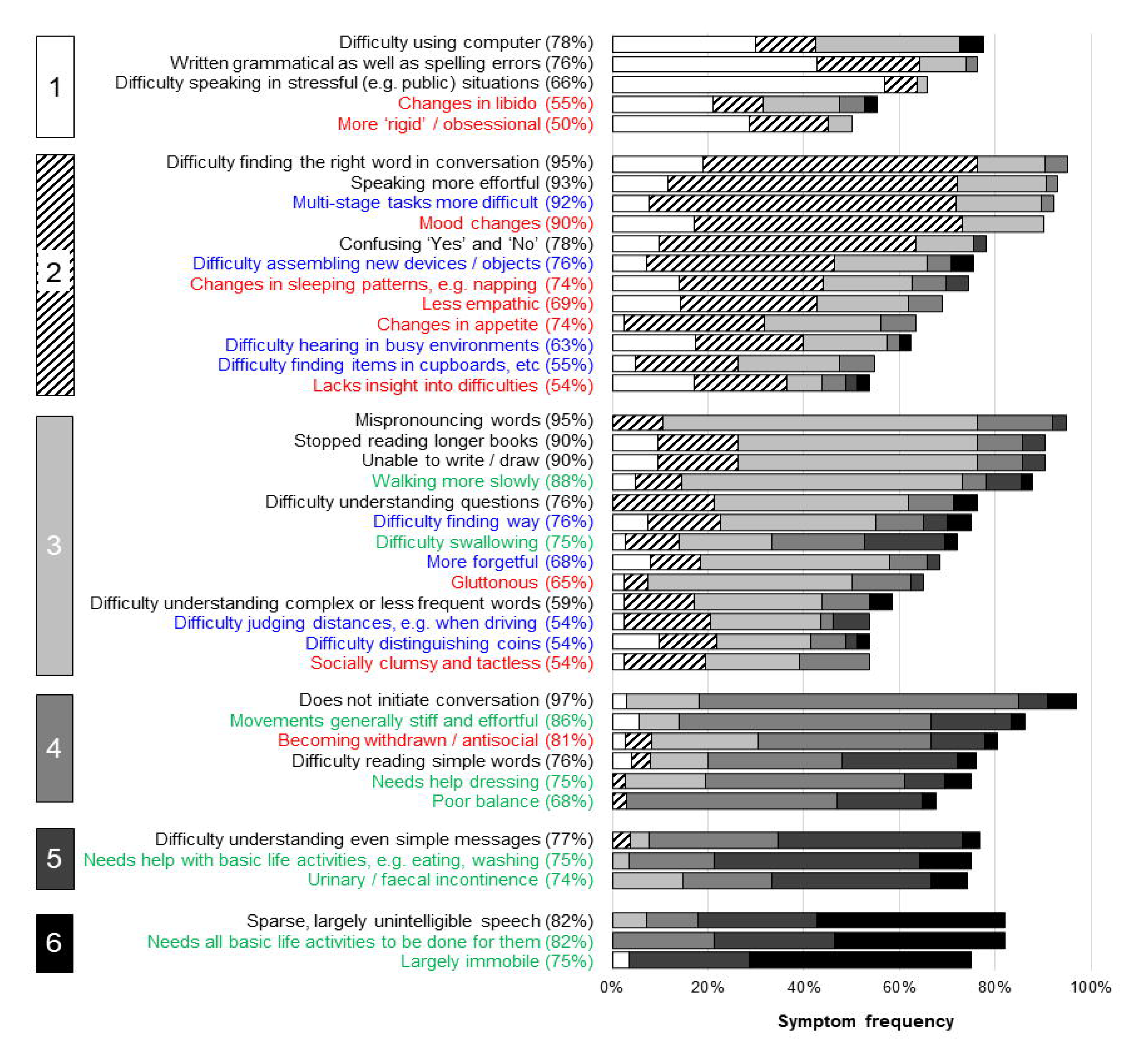
Clinical milestones of primary progressive aphasia evolution. The Figure summarises milestone symptoms and associated management implications over the clinical course of primary progressive aphasia syndromes, as identified from the caregiver survey. The left-hand panels show symptoms which (in that phase of the illness) are more prominent in some PPA syndromes than others; the middle panels show symptoms common to all syndromes. The right-hand panels represent key implications for management during each phase of the illness, as predicted from these milestone symptoms. Written symptom labels are colour-coded based on domains of verbal communication (black), nonverbal thinking (blue), nonverbal conduct and wellbeing (red) and physical symptoms (green) (see text for details).

### Qualitative analysis of survey responses

The qualitative framework comprised five major themes (Table 3; Table S4 online): (i) impact and experience of symptoms; (ii) illness progression/trajectory; (iii) experience of doing the research; (iv) utility of the stages; (v) suggestions for future development/dissemination. Fifteen subthemes were identified within these major themes, and together themes and subthemes encompassed respondents’ experiences of living with PPA and of the staging survey, and their suggestions for implementation.

**Table 3.**
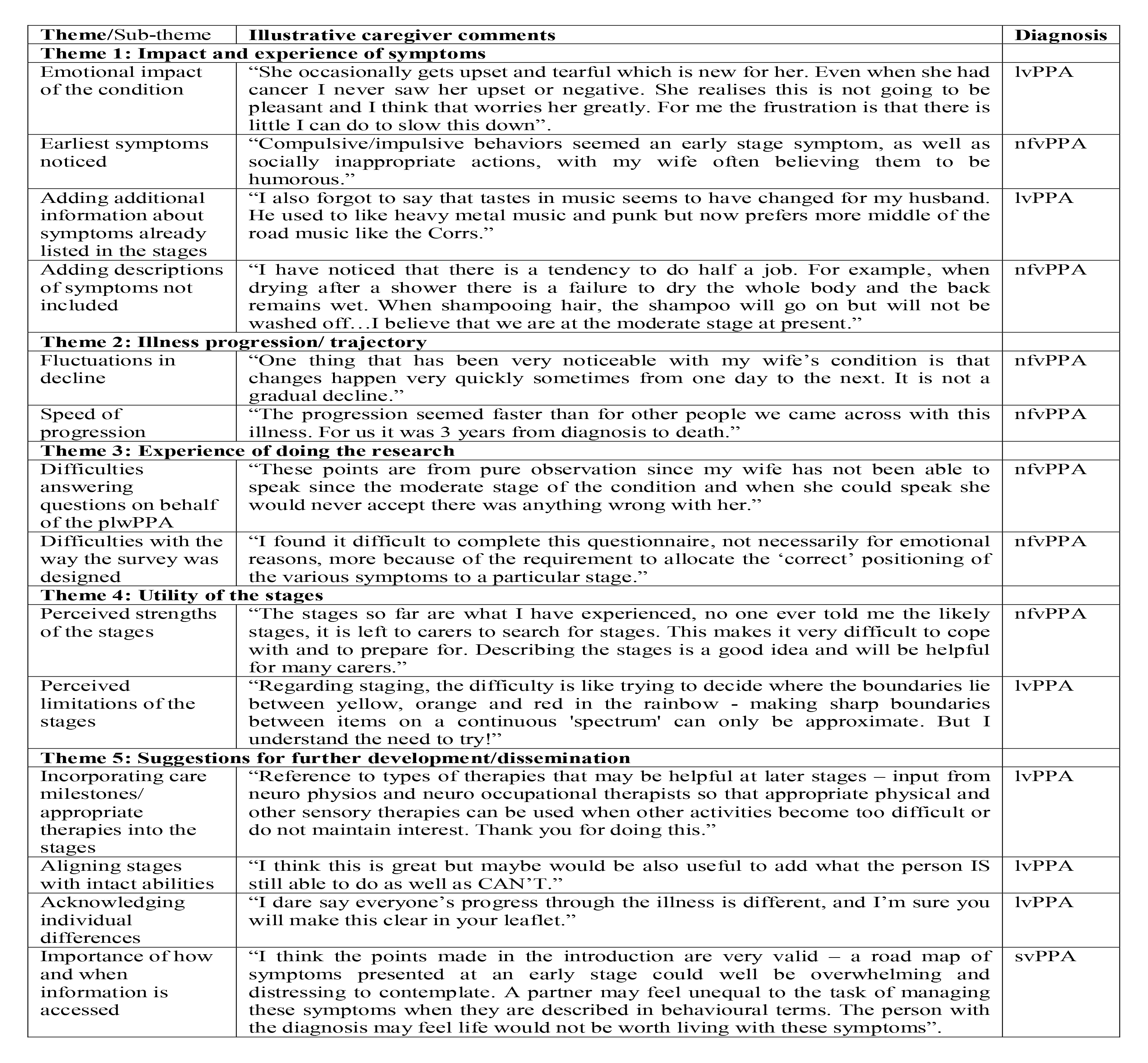
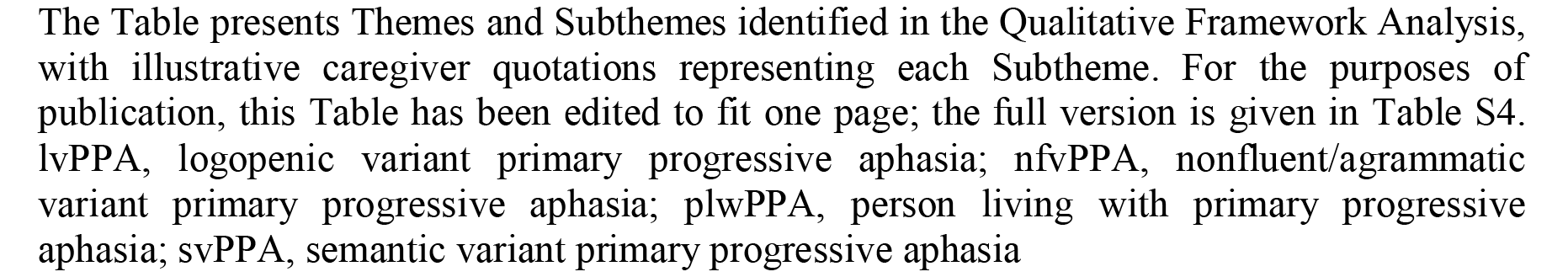
Qualitative framework analysis: themes, sub-themes, and illustrative caregiver comments

## DISCUSSION

Here we propose prototypical, symptom-led clinical stages for each main syndromic variant of PPA, informed by the lived experience of caregivers drawn from a large international research cohort. The proposed staging scheme comprises the information presented in Table 1 and Figures 2-4, which we have collectively termed the PPA Progression Planning Aid (or PPA^2^). This aligns with the structure of the six-stage framework and stage labels (ranging from stage 1, ‘Very mild’ to stage 6, ‘Profound’) used in the Frontotemporal Dementia Rating Scale^14^. Confidence in symptom staging (i.e. the proportion of survey respondents who agreed that a given symptom occurred at a specific stage) varied within and across stages and syndromes: overall, consensus was highest for svPPA but only moderate for nfvPPA, and incomplete for all syndromes and stages. Our survey data suggest that distinct harbinger symptoms of impaired communication function signal the onset of particular PPA syndromes but that nonverbal features are also common from an early stage in all syndromes. The data endorse previous clinical formulations emphasising that PPA variants converge clinically and physical neurological impairments affecting mobility and autonomic function become more salient over time^6, 11^. Hearing changes occurred early in all syndromes, underlining that these language-led dementias are pervasive disorders of communication with important ‘input’ as well as ‘output’ dimensions. Across PPA subtypes, and corroborating findings from previous studies^4, 5^, survey respondents reported an average interval of 2.3 to 3.3 years between symptom onset and diagnosis.

Our findings paint a complex picture of PPA clinical phenotypes. By definition each of the canonical subtypes is led by speech and/or language impairments, and these do show syndromic specificity – broadly, svPPA is led by symptoms consequent to loss of vocabulary and word meaning, while nfvPPA and lvPPA are led by dysfluency that mars conversational discourse and tends to manifest first in situations where there is a ‘performance’ requirement such as speaking in public. Certain features – such as compiling personal ‘dictionaries’ in svPPA and binary reversals in nfvPPA – emerged as leading hallmarks of particular syndromes. In contrast, spelling errors developed early in all syndromes: a potentially useful clinical signal of PPA cases within the much wider spectrum of patients with ‘word finding difficulty’ presenting to memory clinics. Within the diverse repertoire of nonverbal functional abnormalities endorsed as cross-syndromic early features - including difficulty managing devices and sequential tasks more generally, difficulty hearing in busy environments, mood changes and altered socio-emotional reactivity toward other people – there was some evidence for syndromic specificity. Thus, hearing and navigational problems developed earlier in lvPPA than other syndromes, while earliest socio-emotional behavioural alterations took somewhat different forms in particular syndromes (appetite changes in svPPA, mental rigidity in nfvPPA, social withdrawal in lvPPA), consistent with previous work^7, 46–54^. Even at later stages, as increasing difficulties with skilled tasks such as driving and dressing and declining mobility supervened across syndromes, the imprint of particular syndromes was still evident: swallowing and mobility problems developed earlier in nfvPPA than other syndromes, while difficulty recognising familiar people and household items characterised svPPA and visuospatial symptoms were more prominent in lvPPA.

Grouping symptoms by stage suggested functional milestones in the evolution of PPA syndromes (Figure 5): key deficits that predict the sequence of major daily life impacts and associated management needs, through early-, mid and late-phase disease. Both syndrome-specific and cross-syndromic deficits feed into the milestones that characterise each phase of the illness. In early phase disease, loss of communication facility coupled with hearing changes, difficulties with device use and socio-emotional behavioural changes together predict major impacts on occupational and/or social functioning. In mid-phase disease, accumulating language impairments coupled with loss of manual skills and declining mobility predict an increasing need for external carer support. In late-phase disease, inability to communicate coupled with dependency in basic life activities including toileting and deteriorating swallowing function will often prompt a transition to institutional and end-of-life care.

The staging scheme we propose highlights a number of clinical issues with important implications for diagnosis, decisions around support and care needs, and interventions. More reliable detection of the earliest features of PPA would facilitate timelier and more accurate diagnosis of PPA syndromes, and this is particularly apposite given the frequently lengthy delay between symptom onset and diagnosis reported by caregivers (Table 2). The basis for this delay needs elucidation – thus, while medical advice tended on average to be sought earlier in dysfluent syndromes than svPPA (which might reflect the relative salience of speech errors in these syndromes), this varied widely in all syndromes, implying that other factors (perhaps educational, occupational, cultural and/or socioeconomic) importantly influenced the decision. Our findings draw attention to early features of PPA syndromes that do not form part of current consensus diagnostic criteria^1^ – these include both verbal phenomena (such as binary reversals^55^), impaired hearing under challenging listening conditions and behavioural changes affecting libido, sleep, empathy and insight. The prevalence of nonverbal features across PPA syndromes in stage 1 presents a challenge for diagnostic criteria stipulating that language deficits should dominate ‘the initial phases’ of the illness^1^. Equally, the later stages here reveal the landscape of advanced PPA, for which we presently lack tools for the comprehensive assessment of needs and evidence-based interventions. The symptom sequences shown in Figures 2-4 and Tables S1-S3 would enable better clinical prognostication in individual patients, and also allow speech and language therapy and behavioural interventions to be rationally developed, customised and targeted at specific disease subtypes and stages. Current evidence indicates that speech and language interventions that are delivered earlier are more likely to result in maintenance of communication^56^, yet therapists report patients are often referred too late to benefit from such approaches^57^. Mapping individualised trajectories is crucial in light of the widely variable rates at which patients with PPA progress through the illness, an issue endorsed by caregivers in the free-text qualitative data provided (Table 3; Table S4) and fundamental in defining care needs and priorities, and formal statutory care assessments. This information potentially has utility not only for clinicians with experience of PPA, but for allied health multidisciplinary teams who may not always have expert knowledge of PPA to support their treatment planning and service delivery, as well as those with lived experience of the diseases.

From a neurobiological perspective, the staging framework emphasises the ‘trans-syndromic’ nature of canonical PPA variants. This is in accord with recent work emphasising multidimensional phenotypic overlap in the PPA spectrum, arising from the distributed neural network alterations integral to these diseases^36, 58–61^. However, the present findings also underline the importance of the temporal dimension in interpreting the phenotypic spectrum in particular PPA syndromes. In svPPA, symptoms reflect involvement of the dominant followed by the right temporal lobes and their inferior frontal and limbic connections: the ‘semantic appraisal’ network targeted by the proteinopathy in svPPA supports a broad diversity of cognitive and behavioural functions^62–65^, while unambiguously ‘extra-temporal’ (e.g., motor) symptoms occur late. By contrast, in nfvPPA the sequence of multidimensional symptoms suggests relatively early involvement of motor control and basal ganglia networks, consistent with neuroanatomical studies of this syndrome^18, 50^; while in lvPPA, the symptom sequence tracks disease evolution through parietal cortices^66, 67^ and highlights the substantial phenotypic overlap between lvPPA and PCA^2, 22, 36, 58^. The varying ‘confidence’ in symptom staging between PPA syndromes is also informative (Figures 2 to 4; Tables S1 to S3): across stages, consensus was higher for svPPA than other variants, endorsing the view from previous neuropsychological, neuroanatomical and neuropathological studies that this is a highly coherent syndrome, whereas nfvPPA and lvPPA are intrinsically heterogeneous and likely to encompass various sub-syndromes^3, 8, 58,68–70^

While our symptom-led survey methodology builds on recent initiatives toward online collection of large clinical and neuropsychological datasets^71, 72^, its advantages are offset by several limitations. Most obviously, caregiver reports (and indeed the individual diagnoses provided) were retrospective and generally not objectively verifiable. The majority of reports concerned patients in later stages of the illness (Table 2): a retrospective stance is not necessarily a disadvantage (since subtle symptoms may not initially be recognised as such) but it is open to recall bias. On the other hand, most respondents were also reporting on incomplete illness trajectories. Stage divisions and the criteria we applied for symptom inclusion (for example, that a symptom assigned to a given stage should be exhibited by over half of patients moving through that stage) are to some extent arbitrary, and motivated largely by anticipated clinical utility: had alternative criteria been applied, the staging landscape would look rather different, albeit at the expense of sensitivity and/or stage and syndrome specificity. Certain symptoms might in principle be highly specific for particular PPA syndromes while nevertheless developing infrequently (e.g. musicophilia in svPPA). Symptoms were not quantified in frequency or severity, and the symptom descriptors themselves were often broad: ‘difficulty using a computer’, for instance, is likely underpinned by different deficits in particular PPA syndromes and might be further dissected. This caveat may be particularly relevant to socio-emotional behavioural changes such as ‘mental rigidity’ or ‘lack of empathy’ which might conceivably reflect reduced verbal facility manifesting as stereotypies or adynamia. It is not clear how the proposed stages map onto objective measures of disease progression in PPA, or whether a six-stage scheme is optimal; fewer stages would provide greater uniformity within syndromes but lose granularity, while it may be possible in future to adapt the scheme to include a pre-symptomatic ‘Stage 0’ [no cognitive decline] to align with instruments such as the Global Deterioration Rating Scale^13^. The present staging scheme lacks information on the duration of particular stages or the overall tempo of the illness. Clinical experience suggests that individual temporal variability is likely to be wide, a point underlined by the qualitative analysis here (see Table 3; Table S4).

Further, the surveyed cohort here does not represent the full spectrum of PPA. Respondents were all members of PPA support groups who had engaged with clinical diagnostic services, and all were English speakers with internet access to the survey and sufficient technological skills to complete it. Similar limitations apply to most previously published studies of PPA. It is not clear to what extent the proposed staging scheme would be recapitulated in more linguistically and/or socio-culturally diverse populations. Even within the surveyed cohort, the qualitative responses provided by caregivers highlighted the variation in their opinion of the utility, relevance and impact of the approach, based on their personal experience of the illness (see Table 3; Table S4). Moreover, we have focussed here on the three canonical syndromes of PPA: a substantial minority of patients with PPA do not meet consensus criteria for one of these syndromes^21^, and it remains unclear how well such cases fit with the staging scheme.

These caveats suggest directions for future work. The findings should be corroborated, extended and validated in larger and more diverse patient cohorts, including atypical or less common PPA cases beyond the canonical three syndromes and including comparator disease groups to evaluate the specificity of the stages. Prospective, longitudinal studies are required to ensure complete stage coverage without recall bias and to define the temporal dynamics of stage transitions in individual patients. Currently broad symptom categories could be deconstructed and more fine-grained information collected about daily life impact, particularly with a view to consolidating illness milestones. Ideally, first-person patient perspectives would be gathered alongside those of caregivers, to capture experiential features (such as hallucinations and other perceptual disturbances) that tend to elude third-person recording, and to better characterise loss of insight. As with most dementia scales, our staging scheme presently focuses on deficit and disability; there is an opportunity here to contextualise this record of losses by documenting retained capacities that could support interventions, another theme that emerged in qualitative analysis (Table 3; Table S4). This should be informed by an ongoing dialogue both with lay stakeholders and with clinicians, to critique the stages and feed back about their acceptability and utility in diagnosis and care planning. Support groups such as those run by Rare Dementia Support in the UK (www.raredementiasupport.org) will have a central role to play in managing patient and wider public engagement in the further development and translation of the proposed staging framework^73^. Any staging scheme for PPA should be anchored in disease biology and objective biomarkers of disease evolution – symptom led stages should be correlated and elaborated with neuropsychological scores, structural and functional neuroanatomy and extant rating scales for these diseases^14, 28, 63, 74^, as well as CSF and other laboratory indices^75^. Development of quantitative symptom severity scales would facilitate computational modelling approaches, which have been used successfully for tracking of progression in a number of neurodegenerative diseases^53, 54^. Following patients through to post-mortem would allow for relation of clinical symptoms to underlying neuropathology, and genetic cohorts, whilst rare, represent a unique and important opportunity to unpack the very earliest stages of disease (i.e., the putative ‘stage 0’). Application and interpretation of staging would be enhanced by more clinical and demographic detail concerning, factors that modulate the expression of PPA such as developmental dyslexia and educational attainment^11, 76^.

The present findings provide a prototype for the development of a clinically informed staging system for a PPA Progression Planning Aid (PPA^2^). We hope that the work will motivate multi-dimensional, international collaborations to take the essential (and challenging) next steps toward validation. In this spirit, we have included a Spanish translation of Table 1 (“Cross-syndromic descriptions of the PPA Progression Planning Aid (PPA^2^) Stages”) in Supplementary Table S5. The PPA^2^ will also be freely available on the Rare Dementia Support website for caregivers and people with PPA to access, and the succinct symptom labels included in Figures 2-4 and Tables S1-S3 have been designed to be ‘jargon-free’ in the hope that these will have maximal utility for people with different backgrounds and expertise. Our findings argue for a fresh consensus on diagnostic criteria and management guidelines for PPA, to take account of important emerging themes in the clinical phenotyping of these patients, and highlight the importance of learning from people with lived experience of these conditions, in conjunction with clinical and research-derived observations. Examples signalled here include syndromic hallmarks such as early binary reversals in nfvPPA and the prominence of auditory and nonverbal behavioural features across PPA syndromes. A staging system for PPA could provide a bridge to the development of bespoke functional scales of daily life communication^77^ that could in turn serve as outcome measures for clinical trials in these syndromes and language-led AD clinical phenotypes^78^. Looking forward, we envisage that particular symptoms identified as harbingers of early-stage PPA might inform the development of diagnostic ‘cognitive stress tests’ based on the analysis of patients’ spontaneous conversation^79–81^ or requiring production and/or understanding of speech in multi-talker environments or other demanding conditions^52, 82^. However, the overriding value of a PPA staging system, as attested to in the qualitative analysis, will lie in the personalised care of individual patients and families navigating these illnesses – to help them signpost their journey, harness all appropriate supports and treatments, and plan for the future.

## ACKNOWLEDGMENTS

We thank all caregiver respondents for their participation.

