## Supplementary materials for "Symptom-led staging for primary progressive aphasia"

**SUPPLEMENTARY MATERIAL for “Symptom-led staging for primary progressive aphasia”, by CJD Hardy et al**

**Supplementary Table S1.** Summary of survey responses from caregivers for people with svPPA

| **STAGE 1: Very Mild svPPA** | | | | | | | | | | | |
| --- | --- | --- | --- | --- | --- | --- | --- | --- | --- | --- | --- |
| **Original symptom descriptor** | **N responses** | **% present** | **% correct [S1]** | **% S2** | **%S3** | **%S4** | **%S5** | **%S6** | **Action** | **Succinct item wording** | **Neurological interpretation** |
| Problems remembering certain words that are highly specific to the person's interests or hobbies, e.g. a keen gardener might first notice that they are having difficulties with the names for flowers | 27 | 100.0 | **77.8** | 22.2 | 0.0 | 0.0 | 0.0 | 0.0 |  | Difficulty remembering specific familiar (e.g., occupational) vocabulary | Anomia |
| Mood changes, such as becoming more irritable or anxious | 27 | 88.9 | **66.7** | 8.3 | 20.8 | 4.2 | 0.0 | 0.0 |  | Mood changes | Affective alterations |
| Subtle personality changes - e.g. somebody who was previously very careful with money might buy something expensive and out of character | 27 | 63.0 | **58.8** | 17.7 | 17.7 | 5.9 | 0.0 | 0.0 |  | Subtle personality changes | Disinhibition |
| Changes in food preferences or appetite | 25 | 76.0 | **52.6** | 36.8 | 10.5 | 0.0 | 0.0 | 0.0 |  | Changes in appetite | Abnormal eating behaviour |
| Change in libido – an increase or decrease in sexual desire | 25 | 64.0 | **50.0** | 50.0 | 0.0 | 0.0 | 0.0 | 0.0 | Moved to S1 from S2 | Change in libido | Socio-emotional dysfunction |
| Pronunciation errors when reading aloud, especially for unusual 'irregular' words that don't follow regular patterns of pronunciation. For instance, the person may read the word aunt" as "ornt" | 26 | 80.8 | **47.6** | 38.1 | 14.3 | 0.0 | 0.0 | 0.0 |  | Pronunciation errors when reading aloud irregular words (e.g. 'island') | Surface dyslexia |
| Less confidence when engaging in conversations, especially if on the telephone or in groups of people | 27 | 88.9 | **45.8** | 29.2 | 25.0 | 0.0 | 0.0 | 0.0 |  | Less confident engaging in conversations | Impaired discourse |
| Spelling errors, especially for uncommon irregular words that don't follow regular patterns. For example, the person may write the word ache" as "ake" | 27 | 81.5 | **40.9** | 40.9 | 18.2 | 0.0 | 0.0 | 0.0 |  | Spelling errors on less familiar or irregular words (e.g. 'yacht') | Surface dysgraphia |
| **STAGE 2: Mild svPPA** | | | | | | | | | | | |
| **Original symptom descriptor** | **N responses** | **% present** | **%S1** | **% correct [S2]** | **%S3** | **%S4** | **%S5** | **%S6** | **Action** | **Succinct item wording** | **Neurological interpretation** |
| Complicated tasks with multiple steps that the person previously found easy (e.g. cooking an elaborate meal) become more difficult and take the person longer to complete. | 26 | 96.2 | 4.0 | **92.0** | 4.0 | 0.0 | 0.0 | 0.0 |  | Multi-stage tasks more difficult | Executive dysfunction |
| Less self-control, e.g. when it comes to eating chocolate or drinking alcohol. | 26 | 65.4 | 5.9 | **88.2** | 5.9 | 0.0 | 0.0 | 0.0 |  | Gluttonous |  |
| Making lists (‘dictionaries’) of words to be remembered. | 26 | 61.5 | 12.5 | **81.3** | 6.3 | 0.0 | 0.0 | 0.0 |  | Compiles word lists ('dictionaries') | Verbal agnosia |
| Difficulties with reading – stopping reading longer books and instead preferring shorter articles in newspapers or magazines. | 26 | 96.2 | 8.0 | **80.0** | 12.0 | 0.0 | 0.0 | 0.0 |  | Stops reading longer books | Dominant parietal dysfunction |
| Speech is fluent, but seems to ramble and go around the point, e.g. if describing going up on an escalator, they may say, “the moving stairs that take you up”. | 26 | 92.3 | 0.0 | **79.2** | 12.5 | 8.3 | 0.0 | 0.0 |  | Rambling conversation | Circumlocutions |
| Changes in sleeping patterns, e.g. seeming more tired and napping during the day. | 26 | 84.6 | 18.2 | **77.3** | 0.0 | 4.6 | 0.0 | 0.0 |  | Changes in sleeping patterns, e.g. napping | Disordered sleep |
| No longer interested in or enjoys solving crossword puzzles. | 24 | 79.2 | 15.8 | **73.7** | 5.3 | 5.3 | 0.0 | 0.0 |  | No longer enjoys crosswords | Anomia |
| Little insight into the fact that there is anything wrong. | 26 | 73.1 | 26.3 | **73.7** | 0.0 | 0.0 | 0.0 | 0.0 |  | Lacks insight into difficulties | Anosognosia |
| Repetitive/obsessive behaviours | 26 | 92.3 | 8.3 | **66.7** | 20.8 | 4.2 | 0.0 | 0.0 |  | More ‘rigid’ / obsessional | Obsessionality |
| More socially clumsy in situations that require tactfulness. For instance, seeming oddly cheerful when saying hello to somebody at a sombre event like a funeral, or asking a friend or family member a personal question about their appearance. | 26 | 92.3 | 20.8 | **62.5** | 16.7 | 0.0 | 0.0 | 0.0 |  | Socially clumsy and tactless | Socio-emotional dysfunction |
| *Particular problems hearing in busy environments, e.g. a noisy room or a dinner party. | 21 | 71.4 | 26.7 | **53.3** | 13.3 | 6.7 | 0.0 | 0.0 | Added to S2 | Difficulty hearing in busy environments | Central auditory dysfunction |
| †Problems using a computer, e.g being slower to type or enter information | 19 | 84.2 | 12.5 | **43.8** | 43.8 | 0.0 | 0.0 | 0.0 | Added to S2 | Difficulty using computer | Activities of daily living |
| Problems understanding the meaning of more complex or less frequent words | 27 | 85.2 | 26.1 | **43.5** | 30.4 | 0.0 | 0.0 | 0.0 | Moved to S2 from S1 | Difficulty understanding complex or less frequent words | Verbal agnosia |
| †Difficulty assembling new devices/objects | 20 | 60.0 | 16.7 | **33.3** | 33.3 | 16.7 | 0.0 | 0.0 | Added to S2 | Difficulty assembling new devices/objects | Apraxia |
| **STAGE 3: Moderate svPPA** | | | | | | | | | | | |
| **Original symptom descriptor** | **N responses** | **% present** | **%S1** | **% S2** | **% correct [S3]** | **%S4** | **%S5** | **%S6** | **Action** | **Succinct item wording** | **Neurological interpretation** |
| Loss of even common vocabulary, e.g. when asked to get something from the fridge, the person might reply, “What is a fridge?” | 23 | 82.6 | 0.0 | 5.3 | **84.2** | 10.5 | 0.0 | 0.0 |  | Difficulty understanding more common words | Verbal agnosia |
| Some problems with their memory for places or events. | 25 | 100.0 | 8.0 | 12.0 | **80.0** | 0.0 | 0.0 | 0.0 |  | More forgetful | Impaired episodic memory |
| Using very general words like “whatsit” or “that thing” a lot of the time. | 24 | 91.7 | 0.0 | 22.7 | **77.3** | 0.0 | 0.0 | 0.0 |  | Often uses very general words, e.g. "whatsit" | Anomia |
| Marked difficulties in remembering words and names, e.g. forgetting the name of the person who lives down the road. | 25 | 96.0 | 0.0 | 29.2 | **66.7** | 4.2 | 0.0 | 0.0 |  | Difficulty remembering names of people | Anomia |
| Complaining about pains or feelings in the body that don’t seem to have any physical explanation, e.g. headaches, toothache, or pains and feelings in other body parts; and/ or feeling temperature differently to before. | 23 | 69.6 | 0.0 | 31.3 | **62.5** | 6.3 | 0.0 | 0.0 |  | Bodily complaints with no apparent cause | Sensory dysregulation |
| Starting to dislike certain music or other sounds; and/ or complaining of tinnitus or a constant ringing in their ears. | 25 | 52.0 | 23.1 | 0.0 | **61.5** | 15.4 | 0.0 | 0.0 |  | Increased sensitivity to sound / tinnitus | Central auditory dysfunction |
| Finding their way, especially in new places, more difficult. | 24 | 75.0 | 0.0 | 22.2 | **61.1** | 11.1 | 0.0 | 5.6 |  | Difficulty finding way | Topographical agnosia |
| *Getting Yes and No mixed up, for instance saying “No” when asked if they would like a cup of coffee, but really meaning “Yes”. | 20 | 50 | 0.0 | 20.0 | **60.0** | 20.0 | 0.0 | 0.0 | Added to S3 | Confusing 'Yes' and 'No' | Binary reversals |
| Lacking warmth or empathy | 25 | 92.0 | 13.0 | 30.4 | **56.5** | 0.0 | 0.0 | 0.0 |  | Less empathic | Socio-emotional dysfunction |
| *Difficulties with questions, e.g. understanding when choices are being given. | 19 | 89.5 | 0.0 | 29.4 | **52.9** | 17.7 | 0.0 | 0.0 | Added to S3 | Difficulty understanding questions | Receptive agrammatism |
| Showing a sudden love for a particular kind of music or band. | 24 | 33.3 | 0.0 | 37.5 | 62.5 | 0.0 | 0.0 | 0.0 | Removed - not above 50% threshold |  | Central auditory dysfunction |
| **STAGE 4: Severe svPPA** | | | | | | | | | | | |
| **Original symptom descriptor** | **N responses** | **% present** | **%S1** | **%S2** | **%S3** | **% correct [S4]** | **%S5** | **%S6** | **Action** | **Succinct item wording** | **Neurological interpretation** |
| Moving more slowly than before, e.g. when walking from room to room, or getting up from a chair. | 22 | 72.7 | 0.0 | 0.0 | 12.5 | **87.5** | 0.0 | 0.0 |  | Walking more slowly | Parkinsonism |
| Problems recognising household items, e.g. attempting to use bleach as washing-up liquid. | 21 | 90.5 | 0.0 | 5.3 | 10.5 | **84.2** | 0.0 | 0.0 |  | Difficulty recognising household items | Nonverbal agnosia |
| Becoming more withdrawn, not engaging with other people. | 21 | 85.7 | 0.0 | 0.0 | 16.7 | **83.3** | 0.0 | 0.0 |  | Becoming withdrawn / antisocial | Socio-emotional dysfunction |
| Able to carry out some elements of personal care satisfactorily, but needing assistance with some things, e.g. dressing, shaving. | 21 | 85.7 | 0.0 | 5.6 | 11.1 | **72.2** | 11.1 | 0.0 |  | Needs help dressing | Activities of daily living |
| Problems recognising people continue – whilst before the difficulty may have been remembering a person’s name, they may now struggle to recognise that they have even met the person before. | 21 | 100.0 | 0.0 | 0.0 | 28.6 | **66.7** | 4.8 | 0.0 |  | Difficulty recognising familiar people | Prosopagnosia |
| The person makes grammatical as well as spelling errors in emails and notes, etc. | 22 | 86.4 | 0.0 | 5.3 | 31.6 | **63.2** | 0.0 | 0.0 |  | Written grammatical as well as spelling errors | Dysgraphia, expressive agrammatism |
| Problems understanding complicated or long sentences. | 24 | 100.0 | 0.0 | 0.0 | 37.5 | **62.5** | 0.0 | 0.0 |  | Difficulty understanding longer sentences | Receptive agrammatism |
| The person can still speak coherently, but they now rely on ‘stock’ phrases or stories that they will often repeat regardless of context. | 23 | 91.3 | 0.0 | 4.8 | 42.9 | **52.4** | 0.0 | 0.0 |  | Stock phrases | Verbal stereotypies |
| Feeling unsteady and/or falling. | 12 | 66.7 | 0.0 | 0.0 | 12.5 | **37.5** | 37.5 | 12.5 | Moved from S5 to S4 | Poor balance | Postural instability |
| Needing encouraging/ reminding to use the toilet. | 21 | 42.9 | 0.0 | 0.0 | 0.0 | 100.0 | 0.0 | 0.0 | Removed - not above 50% threshold |  | Bladder / bowel dysregulation |
| **STAGE 5: Very Severe svPPA** | | | | | | | | | | | |
| **Original symptom descriptor** | **N responses** | **% present** | **%S1** | **%S2** | **%S3** | **%S4** | **% correct [S5]** | **%S6** | **Action** | **Succinct item wording** | **Neurological interpretation** |
| Difficulties understanding all but the simplest messages. | 10 | 100.0 | 0.0 | 0.0 | 0.0 | 0.0 | **100.0** | 0.0 |  | Difficulty understanding even simple messages | Receptive agrammatism |
| Requiring substantial help with many basic activities, e.g. eating and washing. | 9 | 88.9 | 0.0 | 0.0 | 0.0 | 0.0 | **100.0** | 0.0 |  | Needs help with basic life activities, e.g. eating, washing | Activities of daily living |
| Movements become slower and stiffer | 9 | 66.7 | 0.0 | 0.0 | 0.0 | 0.0 | **83.3** | 16.7 |  | Movements generally stiff and effortful | Parkinsonism |
| Problems recognising themselves in the mirror. | 10 | 60.0 | 0.0 | 0.0 | 16.7 | 0.0 | **83.3** | 0.0 |  | Difficulty recognising self in mirror | Nonverbal agnosia |
| Speech is sparse and largely unintelligible, and now limited to a few words or sounds that may not make sense. | 10 | 50.0 | 0.0 | 0.0 | 20.0 | 0.0 | **80.0** | 0.0 |  | Sparse, largely unintelligible speech | Mutism |
| Increasingly frequent trouble controlling their bladder or bowels. | 10 | 90.0 | 0.0 | 0.0 | 11.1 | 22.2 | **66.7** | 0.0 |  | Urinary / faecal incontinence | Bladder / bowel dysregulation |
| Difficulties recognising food for what it is, e.g. trying to eat inedible objects or eating unheated frozen food without noticing anything is wrong. | 10 | 60.0 | 0.0 | 0.0 | 0.0 | 33.3 | **66.7** | 0.0 |  | Mouthing inedible items | Nonverbal agnosia |
| Able to read and understand only some if any simple words | 13 | 84.6 | 9.1 | 0.0 | 9.1 | 36.4 | **45.5** | 0.0 |  | Difficulty reading simple words | Alexia |
| Problems with swallowing. | 13 | 84.6 | 0.0 | 9.1 | 18.2 | 18.2 | **36.4** | 18.2 |  | Difficulty swallowing | Dysphagia |
| **STAGE 6: Profound svPPA** | | | | | | | | | | | |
| **Original symptom descriptor** | **N responses** | **% present** | **%S1** | **%S2** | **%S3** | **%S4** | **%S5** | **% correct [S6]** | **Action** | **Succinct item wording** | **Neurological interpretation** |
| Unable to perform any acts of daily living and needing to be washed, dressed, fed, etc. by another person. | 8 | 100.0 | 0.0 | 0.0 | 0.0 | 0.0 | 0.0 | **100.0** |  | Needs all basic life activities to be done for them | Activities of daily living |
| There is now almost no speech at all, but the person may make strange sounds (e.g. teeth grinding / popping noises) or laughing, sometimes inappropriately. | 8 | 87.5 | 0.0 | 0.0 | 0.0 | 0.0 | 0.0 | **100.0** |  | Non-verbal sounds in place of speech | Mutism |
| Unable to control most movements without extreme difficulty. The person is mostly confined to chair or bed. | 8 | 50.0 | 0.0 | 0.0 | 0.0 | 0.0 | 0.0 | **100.0** |  | Largely immobile | Parkinsonism |
| The person can no longer write or draw. | 10 | 90.0 | 0.0 | 0.0 | 0.0 | 33.3 | 11.1 | **55.6** |  | Unable to write / draw | Apraxia |
| **Other PPA items** | | | | | | | | | | | |
| **Original symptom descriptor** | **N responses** | **% present** | **%S1** | **%S2** | **%S3** | **%S4** | **%S5** | **%S6** | **Action** | **Succinct item wording** | **Neurological interpretation** |
| Using made-up words that don’t exist, without seeming to be aware that what they have said doesn’t make sense. | 19 | 47.4 | 0.0 | 22.2 | 55.6 | 11.1 | 11.1 | 0.0 | Not added |  | Neologisms |
| Particular problems when speaking in stressful situations, e.g. at a meeting or giving a presentation | 19 | 36.8 | 42.9 | 28.6 | 14.3 | 14.3 | 0.0 | 0.0 | Not added |  | Impaired discourse |
| Problems with ‘spatial orientation’ emerge – the person may have difficulties judging distances or locating objects. | 19 | 21.1 | 25.0 | 0.0 | 75.0 | 0.0 | 0.0 | 0.0 | Not added |  | Visuospatial dysfunction |
| Hallucinations, e.g. seeing or hearing something that isn’t really there. | 19 | 21.1 | 50.0 | 0.0 | 25.0 | 25.0 | 0.0 | 0.0 | Not added |  | Hallucinations |
| **PCA items** | | | | | | | | | | | |
| **Original symptom descriptor** | **N responses** | **% present** | **%S1** | **%S2** | **%S3** | **%S4** | **%S5** | **%S6** | **Action** | **Succinct item wording** | **Neurological interpretation** |
| Difficulty finding things in a handbag, cupboard, etc. | 21 | 47.6 | 0.0 | 30.0 | 50.0 | 20.0 | 0.0 | 0.0 | Not added |  | Visuospatial dysfunction |
| General ‘clumsiness’ (in people who were not previously clumsy), such as knocking things over (inaccurate reaching out), putting a glass down sideways, etc. | 21 | 38.1 | 12.5 | 25.0 | 50.0 | 0.0 | 12.5 | 0.0 | Not added |  | Apraxia, visuospatial dysfunction |
| Experiencing vertigo or other balance problems | 20 | 30.0 | 0.0 | 16.7 | 33.3 | 16.7 | 16.7 | 16.7 | Not added |  | Postural instability |
| Become confused when handling coins due to difficulties telling them apart | 21 | 28.6 | 0.0 | 16.7 | 50.0 | 33.3 | 0.0 | 0.0 | Not added |  | Dominant parietal dysfunction |
| Mixing up left and right | 20 | 15.0 | 0.0 | 33.3 | 33.3 | 33.3 | 0.0 | 0.0 | Not added |  | Dominant parietal dysfunction |
| Little jerky movements in the fingers, arms, or other parts of the body | 21 | 14.3 | 0.0 | 33.3 | 0.0 | 66.7 | 0.0 | 0.0 | Not added |  | Myocolonus |
| Experiencing delusions, e.g. believing that their caregiver is an imposter | 20 | 10.0 | 0.0 | 0.0 | 50.0 | 50.0 | 0.0 | 0.0 | Not added |  | Delusions |
| Difficulty detecting the edge of pavements, paths and steps etc | 21 | 9.5 | 0.0 | 0.0 | 50.0 | 0.0 | 0.0 | 50.0 | Not added |  | Visuospatial dysfunction |
| Experiencing continued deterioration of sensory functions, e.g. partial or complete loss of response to touch | 21 | 9.5 | 0.0 | 0.0 | 0.0 | 50.0 | 0.0 | 50.0 | Not added |  | Sensory dysregulation |
| Difficulty using stairs because of problems with spatial judgment | 20 | 5.0 | 0.0 | 0.0 | 100.0 | 0.0 | 0.0 | 0.0 | Not added |  | Visuospatial dysfunction |
| Be ‘functionally blind’, requiring support in all visually-guided activities | 20 | 5.0 | 0.0 | 0.0 | 100.0 | 0.0 | 0.0 | 0.0 | Not added |  | Visuospatial dysfunction |
| Experiencing odd visual sensations (e.g. occasional changes or washes of colour in the centre or periphery of their vision) | 21 | 4.8 | 0.0 | 0.0 | 0.0 | 100.0 | 0.0 | 0.0 | Not added |  | Early visual dysfunction |
| Being able to see some things but not others, for instance following things that move (e.g. a ball being thrown) but not being able to find static objects (e.g. a ball lying still on the grass in the garden). | 21 | 4.8 | 0.0 | 0.0 | 0.0 | 100.0 | 0.0 | 0.0 | Not added |  | Visuospatial dysfunction |

The table summarises the survey responses given by caregivers for people they cared for with svPPA. If ≥ 50% of respondents indicated a particular symptom was present but of those, a majority indicated that it should have been assigned to an earlier/later stage, that symptom was reassigned accordingly for the final staging (see main text). Original symptom descriptor, the full wording for each symptom that was presented to respondents in the survey. N responses, total number of respondents for that symptom item; %correct, percentage of respondents reporting that symptom was assigned to the correct stage, %Sn, percentage of respondents reporting that symptom was present at a specific stage. Action, summary of decision as to whether to include symptom in final staging system, and where (if cell is blank, this means the item was retained in the stage it was presented to respondents in). Succinct item wording, reduction of original symptom descriptor, homogenised where possible across syndromes. At the end of the survey, respondents were asked additional questions about symptoms present in other PPA syndromes, and in posterior cortical atrophy (PCA): here, respondents had to indicate whether the symptom was present/ absent, and if present, to assign that symptom to a specific stage. *Indicates that an item from another PPA syndrome was endorsed and incorporated into the relevant Stage; †indicates that a PCA symptom was endorsed and incorporated into the relevant Stage. Items that were not endorsed from these additional lists are given at the bottom of the Table.

**Supplementary Table S2.** Summary of survey responses from caregivers for people with nfvPPA

| **STAGE 1: Very Mild nfvPPA** | | | | | | | | | | | |
| --- | --- | --- | --- | --- | --- | --- | --- | --- | --- | --- | --- |
| **Original symptom descriptor** | **N responses** | **% present** | **% correct [S1]** | **% S2** | **%S3** | **%S4** | **%S5** | **%S6** | **Action** | **Succinct item wording** | **Neurological interpretation** |
| Particular problems when speaking in stressful situations, e.g. at a meeting or giving a presentation | 43 | 72.1 | **74.2** | 22.6 | 3.2 | 0.0 | 0.0 | 0 |  | Difficulty speaking in stressful (e.g. public) situations | Impaired discourse |
| Repetitive/obsessive behaviours | 42 | 50.0 | **57.1** | 33.3 | 9.5 | 0.0 | 0.0 | 0 |  | More 'rigid' / obsessional | Obsessionality |
| The person makes grammatical as well as spelling errors in emails and notes, etc. | 42 | 76.2 | **56.3** | 28.1 | 12.5 | 3.1 | 0.0 | 0 |  | Written grammatical as well as spelling errors | Dysgraphia, expressive agrammatism |
| †Problems using a computer, e.g being slower to type or enter information | 41 | 78.0 | **38.7** | 16.1 | 38.7 | 0.0 | 0.0 | 6.45 | Added to S1 | Difficulty using computer | Activities of daily living |
| *Change in libido – an increase or decrease in sexual desire | 38 | 55.3 | **38.1** | 19.1 | 28.6 | 9.5 | 0.0 | 4.8 | Added to S1 | Change in libido | Socio-emotional dysfunction |
| **STAGE 2: Very Mild nfvPPA** | | | | | | | | | | | |
| **Original symptom descriptor** | **N responses** | **% present** | **%S1** | **% correct [S2]** | **%S3** | **%S4** | **%S5** | **%S6** | **Action** | **Succinct item wording** | **Neurological interpretation** |
| Complicated tasks with multiple steps that the person previously found easy (e.g. cooking an elaborate meal) become more difficult and take the person longer to complete. | 39 | 92.3 | 8.3 | **69.4** | 19.4 | 2.8 | 0.0 | 0.0 |  | Multi-stage tasks more difficult | Executive dysfunction |
| Getting Yes and No mixed up, for instance saying “No” when asked if they would like a cup of coffee, but really meaning “Yes”. | 41 | 78.0 | 12.5 | **68.8** | 15.6 | 0.0 | 3.1 | 0.0 |  | Confusing 'Yes' and 'No' | Binary reversals |
| Speaking seems to require the person to use more effort than before, taking them longer to say things, and making them more tired after speaking. | 43 | 93.0 | 12.5 | **65.0** | 20.0 | 2.5 | 0.0 | 0.0 |  | Speaking more effortful | Speech apraxia |
| Mood changes, such as becoming more irritable or anxious. | 41 | 90.2 | 18.9 | **62.2** | 18.9 | 0.0 | 0.0 | 0.0 |  | Mood changes | Affective alterations |
| Difficulties finding the right word to say in conversations. | 42 | 95.2 | 20.0 | **60.0** | 15.0 | 5.0 | 0.0 | 0.0 |  | Difficulty finding the right word in conversation | Anomia |
| †Difficulty assembling new devices/objects | 41 | 75.6 | 9.7 | **51.6** | 25.8 | 6.5 | 0.0 | 6.5 | Added to S2 | Difficulty assembling new devices / objects | Apraxia |
| Changes in food preferences or appetite | 41 | 63.4 | 3.9 | **46.2** | 38.5 | 11.5 | 0.0 | 0.0 |  | Changes in appetite | Abnormal eating behaviour |
| Lacking warmth or empathy | 42 | 69.0 | 20.7 | **41.4** | 27.6 | 10.3 | 0.0 | 0.0 |  | Less empathic | Socio-emotional dysfunction |
| Changes in sleeping patterns, e.g. seeming more tired and napping during the day. | 43 | 74.4 | 18.8 | **40.6** | 25.0 | 9.4 | 6.3 | 0.0 |  | Changes in sleeping patterns, e.g. napping | Disordered sleep |
| †Difficulty finding things in a handbag, cupboard, etc. | 42 | 54.8 | 8.7 | **39.1** | 39.1 | 13.0 | 0.0 | 0.0 | Added to S2 | Difficulty finding items in cupboards, etc | Visuospatial dysfunction |
| *Little insight into the fact that there is anything wrong. | 41 | 53.7 | 31.8 | **36.4** | 13.6 | 9.1 | 4.6 | 4.6 | Added to S2 | Lacks insight into difficulties | Anosognosia |
| *Particular problems hearing in busy environments, e.g. a noisy room or a dinner party. | 40 | 62.5 | 28.0 | **36.0** | 28.0 | 4.0 | 0.0 | 4.0 | Added to S2 | Difficulty hearing in busy environments | Central auditory dysfunction |
| **STAGE 3: Moderate nfvPPA** | | | | | | | | | | | |
| **Original symptom descriptor** | **N responses** | **% present** | **%S1** | **%S2** | **% correct [S3]** | **%S4** | **%S5** | **%S6** | **Action** | **Succinct item wording** | **Neurological interpretation** |
| Speech is slow and difficult to understand, and many words are mispronounced. | 38 | 94.7 | 0.0 | 11.1 | **69.4** | 16.7 | 2.8 | 0.0 |  | Mispronouncing words | Speech apraxia |
| Moving more slowly than before, e.g. when walking from room to room, or getting up from a chair. | 41 | 87.8 | 5.6 | 11.1 | **66.7** | 5.6 | 8.3 | 2.8 |  | Walking more slowly | Parkinsonism |
| Less self-control, e.g. when it comes to eating chocolate or drinking alcohol. | 40 | 65.0 | 3.9 | 7.7 | **65.4** | 19.2 | 3.9 | 0.0 |  | Gluttonous | Abnormal eating behaviour |
| Some problems with their memory for places or events. | 38 | 68.4 | 11.5 | 15.4 | **57.7** | 11.5 | 3.9 | 0.0 |  | More forgetful | Episodic memory |
| Difficulties with reading – stopping reading longer books and instead preferring shorter articles in newspapers or magazines. | 42 | 90.5 | 10.5 | 18.4 | **55.3** | 10.5 | 5.3 | 0.0 |  | Stops reading longer books | Dominant parietal dysfunction |
| *Difficulties with questions, e.g. understanding when choices are being given. | 42 | 76.2 | 0.0 | 28.1 | **53.1** | 12.5 | 0.0 | 6.3 | Added to S3 | Difficulty understanding questions | Receptive agrammatism |
| The person can no longer write or draw. | 30 | 86.7 | 0.0 | 0.0 | **46.2** | 19.2 | 23.1 | 11.5 | Moved to S3 from S5 | Unable to write / draw | Apraxia |
| *Problems understanding the meaning of more complex or less frequent words | 41 | 58.5 | 4.2 | 25.0 | **45.8** | 16.7 | 0.0 | 8.3 | Added to S3 | Difficulty understanding complex or less frequent words | Verbal agnosia |
| *Finding their way, especially in new places, more difficult. | 40 | 75.0 | 10.0 | 20.0 | **43.3** | 13.3 | 6.7 | 6.7 | Added to S3 | Difficulty finding way | Topographical agnosia |
| *Problems with ‘spatial orientation’ emerge – the person may have difficulties judging distances or locating objects. | 39 | 53.8 | 4.8 | 33.3 | **42.9** | 4.8 | 14.3 | 0.0 | Added to S3 | Difficulty judging distances, e.g. when driving | Visuospatial dysfunction |
| †Become confused when handling coins due to difficulties telling them apart | 41 | 53.7 | 18.2 | 22.7 | **36.4** | 13.6 | 4.5 | 4.5 | Added to S3 | Difficulty distinguishing coins | Dominant parietal dysfunction |
| More socially clumsy in situations that require tactfulness. For instance, seeming oddly cheerful when saying hello to somebody at a sombre event like a funeral, or asking a friend or family member a personal question about their appearance. | 41 | 53.7 | 4.6 | 31.8 | **36.4** | 27.3 | 0.0 | 0.0 |  | Socially clumsy and tactless | Socio-emotional dysfunction |
| Problems with swallowing. | 36 | 72.2 | 3.9 | 15.4 | **26.9** | 26.9 | 23.1 | 3.9 | Moved to S3 from S4 | Difficulty swallowing | Dysphagia |
| Showing increased preference for things to be done in a certain way/ at a certain time. | 37 | 48.6 | 5.6 | 16.7 | 61.1 | 11.1 | 5.6 | 0.0 | Removed - not above 50% threshold |  | Obsessionality |
| **STAGE 4: Severe nfvPPA** | | | | | | | | | | | |
| **Original symptom descriptor** | **N responses** | **% present** | **%S1** | **%S2** | **%S3** | **% correct [S4]** | **%S5** | **%S6** | **Action** | **Succinct item wording** | **Neurological interpretation** |
| When the person tries to start a conversation, their speech is usually limited to just a few words; if talking to somebody else, the person may ‘latch on’ to a word that person has said and repeat it. | 33 | 97.0 | 3.1 | 0.0 | 15.6 | **68.8** | 6.3 | 6.3 |  | Does not initiate conversation | Adynamia |
| Feeling unsteady and/or falling | 34 | 67.6 | 0.0 | 4.4 | 0.0 | **65.2** | 26.1 | 4.4 |  | Poor balance | Postural instability |
| Movements become slower and stiffer. | 36 | 86.1 | 6.5 | 0.0 | 9.7 | **61.3** | 19.4 | 3.2 |  | Movements generally stiff and effortful | Parkinsonism |
| Able to carry out some elements of personal care satisfactorily, but needing assistance with some things, e.g. dressing, shaving. | 36 | 75.0 | 0.0 | 3.7 | 22.2 | **55.6** | 11.1 | 7.4 |  | Needs help dressing | Activities of daily living |
| Becoming more withdrawn, not engaging with other people. | 36 | 80.6 | 3.5 | 6.9 | 27.6 | **44.8** | 13.8 | 3.5 |  | Becoming withdrawn / antisocial | Socio-emotional dysfunction |
| Able to read and understand only some if any simple words | 25 | 76.0 | 5.3 | 5.3 | 15.8 | **36.8** | 31.6 | 5.3 | Moved to S4 from S5 | Difficulty reading simple words | Alexia |
| Needing reminding/ encouraging to use the toilet. | 33 | 39.4 | 0.0 | 0.0 | 46.2 | 46.2 | 0.0 | 7.7 | Removed - not above 50% threshold |  | Bladder / bowel dysregulation |
| Hallucinations, e.g. seeing or hearing something that isn’t really there. | 34 | 17.6 | 0.0 | 16.7 | 50.0 | 33.3 | 0.0 | 0.0 | Removed - not above 50% threshold |  | Hallucinations |
| **STAGE 5: Very severe nfvPPA** | | | | | | | | | | | |
| **Original symptom descriptor** | **N responses** | **% present** | **%S1** | **%S2** | **%S3** | **%S4** | **% correct [S5]** | **%S6** | **Action** | **Succinct item wording** | **Neurological interpretation** |
| Requiring substantial help with many basic activities, e.g. eating and washing. | 28 | 75.0 | 0.0 | 0.0 | 4.8 | 23.8 | **57.1** | 14.3 |  | Needs help with basic life activities, e.g. eating, washing | Activities of daily living |
| Difficulties understanding all but the simplest messages. | 26 | 76.9 | 0.0 | 5.0 | 5.0 | 35.0 | **50.0** | 5.0 |  | Difficulty understanding even simple messages | Receptive agrammatism |
| Increasingly frequent trouble controlling their bladder or bowels. | 27 | 74.1 | 0.0 | 0.0 | 20.0 | 25.0 | **45.0** | 10.0 |  | Urinary / faecal incontinence | Bladder / bowel dysregulation |
| There is now almost no speech at all, but the person may make strange sounds (e.g. teeth grinding / popping noises) or laughing, sometimes inappropriately. | 26 | 42.3 | 0.0 | 27.3 | 0.0 | 27.3 | 36.4 | 9.1 | Removed - not above 50% threshold |  | Mutism |
| **STAGE 6: Severe nfvPPA** | | | | | | | | | | | |
| **Original symptom descriptor** | **N responses** | **% present** | **%S1** | **%S2** | **%S3** | **%S4** | **%S5** | **% correct [S6]** | **Action** | **Succinct item wording** | **Neurological interpretation** |
| Unable to control most movements without extreme difficulty. The person is mostly confined to chair or bed. | 28 | 75.0 | 4.8 | 0.0 | 0.0 | 0.0 | 33.3 | **61.9** |  | Largely immobile | Parkinsonism |
| Speech is sparse and largely unintelligible, and now limited to a few words or sounds that may not make sense. | 28 | 82.1 | 0.0 | 0.0 | 8.7 | 13.0 | 30.4 | **47.8** |  | Sparse, largely unintelligible speech | Mutism |
| Unable to perform any acts of daily living and needing to be washed, dressed, fed, etc. by another person. | 28 | 82.1 | 0.0 | 0.0 | 0.0 | 26.1 | 30.4 | **43.5** |  | Needs all basic life activities to be done for them | Activities of daily living |
| **Other PPA items** | | | | | | | | | | | |
| **Original symptom descriptor** | **N responses** | **% present** | **%S1** | **%S2** | **%S3** | **%S4** | **%S5** | **%S6** | **Action** | **Succinct item wording** | **Neurological interpretation** |
| Problems recognising household items, e.g. attempting to use bleach as washing-up liquid. | 41 | 43.9 | 0.0 | 27.8 | 38.9 | 27.8 | 0.0 | 5.56 | Not added |  | Nonverbal agnosia |
| Problems recognising people. | 41 | 41.5 | 12.5 | 6.3 | 43.8 | 18.8 | 6.25 | 12.5 | Not added |  | Prosopagnosia |
| Complaining about pains or feelings in the body that don’t seem to have an easy physical explanation, e.g. headaches, toothache, or pains and feelings in other body parts; and/ or feeling temperature differently to before. | 41 | 41.5 | 11.8 | 41.2 | 41.2 | 5.9 | 0.0 | 0.0 | Not added |  | Sensory dysregulation |
| Starting to dislike certain music or sounds; and/ or complaining of tinnitus or a constant ringing in their ears. | 41 | 29.3 | 33.3 | 8.3 | 41.7 | 16.7 | 0.0 | 0.0 | Not added |  | Central auditory dysfunction |
| Showing a sudden love for a particular kind of music or band. | 41 | 14.6 | 0.0 | 16.7 | 66.7 | 0.0 | 0 | 16.67 | Not added |  | Central auditory dysfunction |
| **PCA items** | | | | | | | | | | | |
| **Original symptom descriptor** | **N responses** | **% present** | **%S1** | **%S2** | **%S3** | **%S4** | **%S5** | **%S6** | **Action** | **Succinct item wording** | **Neurological interpretation** |
| Experiencing vertigo or other balance problems | 41 | 48.8 | 25.0 | 15.0 | 40.0 | 15.0 | 0.0 | 5.0 | Not added |  | Postural instability |
| Difficulty detecting the edge of pavements, paths and steps etc | 41 | 46.3 | 0.0 | 21.1 | 36.8 | 15.8 | 26.3 | 0.0 | Not added |  | Visuospatial dysfunction |
| Little jerky movements in the fingers, arms, or other parts of the body | 42 | 42.9 | 22.2 | 11.1 | 33.3 | 22.2 | 11.1 | 0.0 | Not added |  | Myocolonus |
| Mixing up left and right | 41 | 39.0 | 6.3 | 56.3 | 25.0 | 0.0 | 12.5 | 0.0 | Not added |  | Dominant parietal dysfunction |
| Difficulty using stairs because of problems with spatial judgment | 42 | 38.1 | 12.5 | 12.5 | 31.3 | 25.0 | 12.5 | 6.3 | Not added |  | Visuospatial dysfunction |
| General ‘clumsiness’ (in people who were not previously clumsy), such as knocking things over (inaccurate reaching out), putting a glass down sideways, etc. | 42 | 35.7 | 26.7 | 20.0 | 40.0 | 13.3 | 0.0 | 0.0 | Not added |  | Apraxia, visuospatial dysfunction |
| Experiencing continued deterioration of sensory functions, e.g. partial or complete loss of response to touch | 39 | 28.2 | 9.1 | 18.2 | 18.2 | 36.4 | 0.0 | 18.2 | Not added |  | Sensory dysregulation |
| Being able to see some things but not others, for instance following things that move (e.g. a ball being thrown) but not being able to find static objects (e.g. a ball lying still on the grass in the garden). | 41 | 22.0 | 44.4 | 11.1 | 11.1 | 22.2 | 11.1 | 0.0 | Not added |  | Visuospatial dysfunction |
| Be ‘functionally blind’, requiring support in all visually-guided activities | 41 | 12.2 | 20.0 | 20.0 | 20.0 | 40.0 | 0.0 | 0.0 | Not added |  | Visuospatial dysfunction |
| Experiencing delusions, e.g. believing that their caregiver is an imposter | 42 | 11.9 | 20.0 | 40.0 | 0.0 | 40.0 | 0.0 | 0.0 | Not added |  | Delusions |
| Experiencing odd visual sensations (e.g. occasional changes or washes of colour in the centre or periphery of their vision) | 42 | 7.1 | 33.3 | 0.0 | 66.7 | 0.0 | 0.0 | 0.0 | Not added |  | Early visual dysfunction |

The table summarises the survey responses given by caregivers for people they cared for with nfvPPA. If ≥ 50% of respondents indicated a particular symptom was present but of those, a majority indicated that it should have been assigned to an earlier/later stage, that symptom was reassigned accordingly for the final staging (see main text). Original symptom descriptor, the full wording for each symptom that was presented to respondents in the survey. N responses, total number of respondents for that symptom item; % present, percentage of respondents reporting that symptom as present in the person they care(d) for, at any stage of the disease; % earlier, percentage of respondents reporting that symptom occurred at an earlier stage than assigned; % correct, percentage of respondents reporting that symptom was assigned to the correct stage; % later, percentage of respondents reporting that symptom occurred at a later stage. Action, summary of decision as to whether to include symptom in final staging system, and where (if cell is blank, this means the item was retained in the stage it was presented to respondents in). Succinct item wording, reduction of original symptom descriptor, homogenised where possible across syndromes. At the end of the survey, respondents were asked additional questions about symptoms present in other PPA syndromes, and in posterior cortical atrophy (PCA): here, respondents had to indicate whether the symptom was present/ absent, and if present, to assign that symptom to a specific stage. *Indicates that an item from another PPA syndrome was endorsed and incorporated into the relevant Stage; †indicates that a PCA symptom was endorsed and incorporated into the relevant Stage.

**Supplementary Table S3.** Summary of survey responses from caregivers for people with lvPPA

| **STAGE 1: Very Mild lvPPA** | | | | | | | | | | | |
| --- | --- | --- | --- | --- | --- | --- | --- | --- | --- | --- | --- |
| **Original symptom descriptor** | **N responses** | **% present** | **% correct [S1]** | **% S2** | **%S3** | **%S4** | **%S5** | **%S6** | **Action** | **Succinct item wording** | **Neurological interpretation** |
| Particular problems hearing in busy environments, e.g. a noisy room or a dinner party. | 36 | 63.9 | **87.0** | 13.0 | 0.0 | 0.0 | 0.0 | 0.0 |  | Difficulty hearing in busy environments | Central auditory dysfunction |
| Difficulties finding names for people and things, though it feels to the person as if it’s on the ‘tip of their tongue’. | 36 | 83.3 | **83.3** | 16.7 | 0.0 | 0.0 | 0.0 | 0.0 |  | Difficulty finding names | Anomia |
| Losing the ‘thread’ of sentences, e.g. starting to stay something but then trailing off. | 36 | 77.8 | **75.0** | 17.9 | 7.1 | 0.0 | 0.0 | 0.0 |  | Losing the 'thread' of sentences | Impaired verbal short term memory |
| *Particular problems when speaking in stressful situations, e.g. at a meeting or giving a presentation | 28 | 64.3 | **66.7** | 22.2 | 5.6 | 5.6 | 0.0 | 0.0 | Added to S1 | Difficulty speaking in stressful (e.g. public) situations | Impaired discourse |
| Becoming more withdrawn, not engaging with other people. | 36 | 66.7 | **54.2** | 41.7 | 4.2 | 0.0 | 0.0 | 0.0 |  | Becoming withdrawn / antisocial | Socio-emotional dysfunction |
| **STAGE 2: Mild lvPPA** | | | | | | | | | | | |
| **Original symptom descriptor** | **N responses** | **% present** | **%S1** | **% correct [S2]** | **%S3** | **%S4** | **%S5** | **%S6** | **Action** | **Succinct item wording** | **Neurological interpretation** |
| Getting parts of words mixed up or confused with other words, e.g. saying “staffolding” instead of “scaffolding”, or “aminal” instead of “animal”. | 34 | 50.0 | 17.7 | **76.5** | 5.9 | 0.0 | 0.0 | 0.0 |  | Mispronouncing words | Phonological errors |
| Problems with confusion around numbers, e.g. struggling to work out how to split a bill in a restaurant, or how much change they should be owed. | 34 | 85.3 | 13.8 | **72.4** | 10.3 | 0.0 | 0.0 | 3.5 |  | Difficulty with numerical tasks | Dominant parietal dysfunction |
| Speech often contains pauses as the person searches for the right word to say. | 34 | 94.1 | 12.5 | **71.9** | 15.6 | 0.0 | 0.0 | 0.0 |  | Conversation often contains pauses | Anomia |
| Mood changes, such as becoming more irritable or anxious. | 34 | 79.4 | 18.5 | **70.4** | 11.1 | 0.0 | 0.0 | 0.0 |  | Mood changes | Affective alterations |
| Finding writing more difficult. Friends and family might start to notice spelling mistakes in emails and notes. | 35 | 85.7 | 20.0 | **70.0** | 6.7 | 3.3 | 0.0 | 0.0 |  | Spelling errors | Dysgraphia |
| Some problems with their memory for places or events | 34 | 85.3 | 27.6 | **62.1** | 10.3 | 0.0 | 0.0 | 0.0 |  | More forgetful | Impaired episodic memory |
| †Problems using a computer, e.g being slower to type or enter information | 31 | 83.9 | 11.5 | **61.5** | 26.9 | 0.0 | 0.0 | 0.0 | Added to S2 | Difficulty using computer | Activities of daily living |
| Finding their way, especially in new places, more difficult. | 34 | 82.4 | 28.6 | **60.7** | 10.7 | 0.0 | 0.0 | 0.0 |  | Difficulty finding way | Topographical agnosia |
| †Difficulty assembling new devices/objects | 29 | 65.5 | 5.3 | **57.9** | 31.6 | 5.3 | 0.0 | 0.0 | Added to S2 | Difficulty assembling new devices/objects | Apraxia |
| Loss of pleasure in reading, e.g. somebody who was previously a big reader may start reading less than they did before. | 35 | 77.1 | 29.6 | **51.9** | 14.8 | 3.7 | 0.0 | 0.0 |  | Loss of pleasure in reading | Dominant parietal dysfunction |
| **STAGE 3: Moderate lvPPA** | | | | | | | | | | | |
| **Original symptom descriptor** | **N responses** | **% present** | **%S1** | **%S2** | **% correct [S3]** | **%S4** | **%S5** | **%S6** | **Action** | **Succinct item wording** | **Neurological interpretation** |
| Problems understanding complicated or long sentences. | 31 | 100.0 | 6.5 | 12.9 | **74.2** | 6.5 | 0.0 | 0.0 |  | Difficulty understanding longer sentences | Receptive agrammatism |
| Complicated tasks with multiple steps that the person previously found easy (e.g. cooking an elaborate meal) become more difficult and take the person longer to complete. | 30 | 86.7 | 0.0 | 26.9 | **69.2** | 3.9 | 0.0 | 0.0 |  | Multi-stage tasks more difficult | Executive dysfunction |
| Problems with ‘spatial orientation’ emerge – the person may have difficulties judging distances or locating objects. | 29 | 65.5 | 5.3 | 21.1 | **68.4** | 5.3 | 0.0 | 0.0 |  | Difficulty judging distances, e.g. when driving | Visuospatial dysfunction |
| †Being able to see some things but not others, for instance following things that move (e.g. a ball being thrown) but not being able to find static objects (e.g. a ball lying still on the grass in the garden). | 29 | 51.7 | 6.7 | 20.0 | **53.3** | 20.0 | 0.0 | 0.0 | Added to S3 | Better able to see moving than static objects (or vice versa) | Visuospatial dysfunction |
| †Mixing up left and right | 29 | 55.2 | 25.0 | 18.8 | **43.8** | 12.5 | 0.0 | 0.0 | Added to S3 | Confuses left / right | Dominant parietal dysfunction |
| †Become confused when handling coins due to difficulties telling them apart | 29 | 58.6 | 5.9 | 29.4 | **41.2** | 23.5 | 0.0 | 0.0 | Added to S3 | Difficulty distinguishing coins | Dominant parietal dysfunction |
| †Difficulty finding things in a handbag, cupboard, etc. | 29 | 82.8 | 8.3 | 33.3 | **37.5** | 16.7 | 4.2 | 0.0 | Added to S3 | Difficulty finding items in cupboards, etc | Visuospatial dysfunction |
| The person makes grammatical as well as spelling errors in emails and notes, etc. | 30 | 90.0 | 0.0 | 22.2 | **37.0** | 37.0 | 0.0 | 3.7 | Moved to S3 from S4 | Written grammatical as well as spelling errors | Dysgraphia, expressive agrammatism |
| Repetitive/obsessive behaviours | 31 | 48.4 | 13.3 | 6.7 | 60.0 | 20.0 | 0.0 | 0.0 | Removed - not above 50% threshold |  | Obsessionality |
| **STAGE 4: Severe lvPPA** | | | | | | | | | | | |
| **Original symptom descriptor** | **N responses** | **% present** | **%S1** | **%S2** | **%S3** | **% correct [S4]** | **%S5** | **%S6** | **Action** | **Succinct item wording** | **Neurological interpretation** |
| Able to carry out some elements of personal care satisfactorily, but needing assistance with some things, e.g. dressing, shaving. | 28 | 71.4 | 5.0 | 0.0 | 20.0 | **70.0** | 5.0 | 0.0 |  | Needs help dressing | Activities of daily living |
| Appearing to be more vacant and not as alert as previously. | 29 | 75.9 | 4.6 | 4.6 | 18.2 | **68.2** | 4.6 | 0.0 |  | Sometimes seems 'vacant' | Alertness |
| Pacing up and down/ constantly walking. | 28 | 53.6 | 0.0 | 6.7 | 13.3 | **66.7** | 13.3 | 0.0 |  | Paces restlessly | Akathisia |
| Problems understanding the meaning of more complex or less frequent words | 30 | 80.0 | 0.0 | 8.3 | 29.2 | **62.5** | 0.0 | 0.0 |  | Difficulty understanding complex or less frequent words | Verbal agnosia |
| Difficulties with questions, e.g. understanding when choices are being given. | 31 | 93.5 | 0.0 | 0.0 | 37.9 | **62.1** | 0.0 | 0.0 |  | Difficulty understanding questions | Receptive agrammatism |
| Moving more slowly than before, e.g. when walking from room to room, or getting up from a chair. | 29 | 72.4 | 4.8 | 4.8 | 28.6 | **61.9** | 0.0 | 0.0 |  | Walking more slowly | Parkinsonism |
| Changes in sleeping patterns, e.g. seeming more tired and napping during the day. | 28 | 82.1 | 0.0 | 13.0 | 26.1 | **60.9** | 0.0 | 0.0 |  | Changes in sleeping patterns, e.g. napping | Disordered sleep |
| Getting Yes and No mixed up, for instance saying “No” when asked if they would like a cup of coffee, but really meaning “Yes”. | 31 | 74.2 | 4.4 | 13.0 | 17.4 | **60.9** | 4.4 | 0.0 |  | Confusing 'Yes' and 'No' | Binary reversals |
| The person can no longer write or draw. | 14 | 92.9 | 0.0 | 0.0 | 15.4 | **53.9** | 15.4 | 15.4 | Moved to S4 from S6 | Unable to write / draw | Apraxia |
| Using made-up words that don’t exist, without seeming to be aware that what they have said doesn’t make sense. | 28 | 53.6 | 0.0 | 6.7 | 40.0 | **53.3** | 0.0 | 0.0 |  | Uses some 'made-up' words | Neologisms |
| *Problems recognising people. | 28 | 50.0 | 0.0 | 0.0 | 21.4 | **50.0** | 21.4 | 7.1 | Added to S4 | Difficulty recognising familiar people | Prosopagnosia |
| Needing reminding/ encouraging to use the toilet. | 28 | 39.3 | 0.0 | 0.0 | 9.1 | 81.8 | 9.1 | 0.0 | Removed - not above 50% threshold |  | Bladder / bowel dysregulation |
| **STAGE 5: Very severe lvPPA** | | | | | | | | | | | |
| **Original symptom descriptor** | **N responses** | **% present** | **%S1** | **%S2** | **%S3** | **%S4** | **% correct [S5]** | **%S6** | **Action** | **Succinct item wording** | **Neurological interpretation** |
| Problems with swallowing | 17 | 52.9 | 0.0 | 0.0 | 0.0 | 0.0 | **88.9** | 11.1 |  | Difficulty swallowing | Dysphagia |
| Requiring substantial help with many basic activities, e.g. eating and washing. | 17 | 76.5 | 0.0 | 0.0 | 15.4 | 7.7 | **76.9** | 0.0 |  | Needs help with basic life activities, e.g. eating, washing | Activities of daily living |
| Movements become slower and stiffer. | 17 | 76.5 | 0.0 | 0.0 | 0.0 | 23.1 | **76.9** | 0.0 |  | Movements generally stiff and effortful | Parkinsonism |
| Speech is sparse and largely unintelligible, and now limited to a few words or sounds that may not make sense. | 18 | 88.9 | 0.0 | 0.0 | 6.3 | 18.8 | **75.0** | 0.0 |  | Sparse, largely unintelligible speech | Mutism |
| Difficulties understanding all but the simplest messages. | 18 | 88.9 | 0.0 | 0.0 | 6.3 | 18.8 | **75.0** | 0.0 |  | Difficulty understanding even simple messages | Receptive agrammatism |
| Feeling unsteady and/or falling | 18 | 61.1 | 9.1 | 0.0 | 0.0 | 9.1 | **72.7** | 9.1 |  | Poor balance | Postural instability |
| Increasingly frequent trouble controlling their bladder or bowels. | 16 | 62.5 | 0.0 | 0.0 | 0.0 | 40.0 | **60.0** | 0.0 |  | Urinary / faecal incontinence | Bladder / bowel dysregulation |
| Difficulties with positioning the body in space, e.g. the person may seem to find it hard to work out how to position their body to get into a car. | 17 | 64.7 | 0.0 | 0.0 | 9.1 | 27.3 | **54.5** | 9.1 |  | Difficulty positioning body in space | Visuospatial dysfunction |
| Able to read and understand only some if any simple words | 17 | 82.4 | 0.0 | 0.0 | 0.0 | 50.0 | **42.9** | 7.1 |  | Difficulty reading simple words | Alexia |
| **STAGE 6: Severe lvPPA** | | | | | | | | | | | |
| **Original symptom descriptor** | **N responses** | **% present** | **%S1** | **%S2** | **%S3** | **%S4** | **%S5** | **% correct [S6]** | **Action** | **Succinct item wording** | **Neurological interpretation** |
| Unable to control most movements without extreme difficulty. The person is mostly confined to chair or bed. | 11 | 63.6 | 0.0 | 0.0 | 0.0 | 0.0 | 0.0 | **100.0** |  | Largely immobile | Parkinsonism |
| Unable to perform any acts of daily living and needing to be washed, dressed, fed, etc. by another person. | 11 | 81.8 | 0.0 | 0.0 | 0.0 | 0.0 | 11.1 | **88.9** |  | Needs all basic life activities to be done for them | Activities of daily living |
| There is now almost no speech at all, but the person may make strange sounds (e.g. teeth grinding / popping noises) or laughing, sometimes inappropriately. | 11 | 90.9 | 0.0 | 0.0 | 0.0 | 0.0 | 30.0 | **70.0** |  | Non-verbal sounds in place of speech | Mutism |
| **Other PPA items** | | | | | | | | | | | |
| **Original symptom descriptor** | **N responses** | **% present** | **%S1** | **%S2** | **%S3** | **%S4** | **%S5** | **%S6** | **Action** | **Succinct item wording** | **Neurological interpretation** |
| Changes in food preferences or appetite. | 29 | 41.4 | 8.3 | 16.7 | 33.3 | 16.7 | 8.3 | 16.7 | Not added |  | Abnormal eating behaviour |
| Little insight into the fact that there is anything wrong. | 28 | 39.3 | 18.2 | 18.2 | 54.6 | 9.1 | 0.0 | 0.0 | Not added |  | Anosognosia |
| Hallucinations, e.g. seeing or hearing something that isn’t really there. | 28 | 39.3 | 0.0 | 18.2 | 36.4 | 27.3 | 18.2 | 0.0 | Not added |  | Hallucinations |
| Complaining about pains or feelings in the body that don’t seem to have an easy physical explanation, e.g. headaches, toothache, or pains and feelings in other body parts; and/ or feeling temperature differently to before. | 29 | 37.9 | 9.1 | 9.1 | 63.6 | 18.2 | 0.0 | 0.0 | Not added |  | Sensory dysregulation |
| Change in libido – an increase or decrease in sexual desire | 29 | 34.5 | 60.0 | 10.0 | 20.0 | 10.0 | 0.0 | 0.0 | Not added |  | Socio-emotional dysfunction |
| Problems recognising household items, e.g. attempting to use bleach as washing-up liquid. | 28 | 21.4 | 16.7 | 0.0 | 16.7 | 50.0 | 16.7 | 0.0 | Not added |  | Nonverbal agnosia |
| Starting to dislike certain music or sounds; and/ or complaining of tinnitus or a constant ringing in their ears. | 30 | 20.0 | 16.7 | 16.7 | 33.3 | 33.3 | 0.0 | 0.0 | Not added |  | Central auditory dysfunction |
| Showing a sudden love for a particular kind of music or band. | 28 | 14.3 | 25.0 | 0.0 | 0.0 | 25.0 | 50.0 | 0.0 | Not added |  | Central auditory dysfunction |
| **PCA items** | | | | | | | | | | | |
| **Original symptom descriptor** | **N responses** | **% present** | **%S1** | **%S2** | **%S3** | **%S4** | **%S5** | **%S6** | **Action** | **Succinct item wording** | **Neurological interpretation** |
| Difficulty detecting the edge of pavements, paths and steps etc | 29 | 44.8 | 0.0 | 0.0 | 23.1 | 46.2 | 23.1 | 7.7 | Not added |  | Visuospatial dysfunction |
| General ‘clumsiness’ (in people who were not previously clumsy), such as knocking things over (inaccurate reaching out), putting a glass down sideways, etc. | 28 | 42.9 | 16.7 | 0.0 | 16.7 | 33.3 | 16.7 | 16.7 | Not added |  | Apraxia, visuospatial dysfunction |
| Little jerky movements in the fingers, arms, or other parts of the body | 29 | 41.4 | 16.7 | 0.0 | 8.3 | 58.3 | 16.7 | 0.0 | Not added |  | Myocolonus |
| Experiencing vertigo or other balance problems | 29 | 31.0 | 11.1 | 11.1 | 22.2 | 33.3 | 22.2 | 0.0 | Not added |  | Balance |
| Difficulty using stairs because of problems with spatial judgment | 29 | 31.0 | 0.0 | 0.0 | 22.2 | 66.7 | 0.0 | 11.1 | Not added |  | Visuospatial dysfunction |
| Be ‘functionally blind’, requiring support in all visually-guided activities | 29 | 13.8 | 0.0 | 0.0 | 50.0 | 25.0 | 0.0 | 25.0 | Not added |  | Visuospatial dysfunction |
| Experiencing delusions, e.g. believing that their caregiver is an imposter | 29 | 13.8 | 0.0 | 25.0 | 25.0 | 25.0 | 25.0 | 0.0 | Not added |  | Delusions |
| Experiencing odd visual sensations (e.g. occasional changes or washes of colour in the centre or periphery of their vision) | 29 | 3.4 | 0.0 | 0.0 | 0.0 | 0.0 | 100.0 | 0.0 | Not added |  | Early visual dysfunction |
| Experiencing continued deterioration of sensory functions, e.g. partial or complete loss of response to touch | 29 | 0.0 | 0.0 | 0.0 | 0.0 | 0.0 | 0.0 | 0.0 | Not added |  | Sensory dysregulation |

The table summarises the survey responses given by caregivers for people they cared for with lvPPA. If ≥ 50% of respondents indicated a particular symptom was present but of those, a majority indicated that it should have been assigned to an earlier/later stage, that symptom was reassigned accordingly for the final staging (see main text). Original symptom descriptor, the full wording for each symptom that was presented to respondents in the survey. N responses, total number of respondents for that symptom item; % present, percentage of respondents reporting that symptom as present in the person they care(d) for, at any stage of the disease; % earlier, percentage of respondents reporting that symptom occurred at an earlier stage than assigned; % correct, percentage of respondents reporting that symptom was assigned to the correct stage; % later, percentage of respondents reporting that symptom occurred at a later stage. Action, summary of decision as to whether to include symptom in final staging system, and where (if cell is blank, this means the item was retained in the stage it was presented to respondents in). Succinct item wording, reduction of original symptom descriptor, homogenised where possible across syndromes. At the end of the survey, respondents were asked additional questions about symptoms present in other PPA syndromes, and in posterior cortical atrophy (PCA): here, respondents had to indicate whether the symptom was present/ absent, and if present, to assign that symptom to a specific stage. *Indicates that an item from another PPA syndrome was endorsed and incorporated into the relevant Stage; †indicates that a PCA symptom was endorsed and incorporated into the relevant Stage.

**Supplementary Table S4.** Qualitative framework analysis: themes, sub-themes, and illustrative caregiver comments

| **Theme/**Sub-theme | **Illustrative caregiver comments** | **Diagnosis** |
| --- | --- | --- |
| **Theme 1: Impact and experience of symptoms** | |  |
| Emotional impact of the condition | “She occasionally gets upset and tearful which is new for her. Even when she had cancer I never saw her upset or negative. She realises this is not going to be pleasant and I think that worries her greatly. For me the frustration is that there is little I can do to slow this down”. | lvPPA |
| Earliest symptoms noticed | “Compulsive/impulsive behaviors seemed an early stage symptom, as well as socially inappropriate actions, with my wife often believing them to be humorous.” | nfvPPA |
|  | “The first thing we really noticed was being unable to read a book, used to read 2-3 per week and inability to follow television programmes, as well as using the same word repeatedly in conversation.” | nfvPPA |
| Adding additional information about symptoms already listed in the stages | ”I find it helpful to think about the sort of sentences mum wouldn’t understand (so to give examples what the language difficulties are like since language is the primary issue) when explaining to people. So for example, to think, if I said ‘mum, can you bring me a plate?’ she wouldn’t understand that as a sentence initially, but if she grasped the word plate (either because I point/ on this occasion the word is familiar) she would know what I meant (i.e. the concept of the sentence makes sense to her but she hasn’t computed the meaning of words”. | svPPA |
|  | “I also forgot to say that tastes in music seems to have changed for my husband. He used to like heavy metal music and punk but now prefers more middle of the road music like the Corrs. Perhaps this is because it is less noisy! He has become increasingly sensitive to music in the last few years and now even taps his feet and claps his hands when he hears something he likes on an advert.” | lvPPA |
| Adding descriptions of symptoms not included | “I have noticed that there is a tendency to do half a job. For example, when drying after a shower there is a failure to dry the whole body and the back remains wet. When shampooing hair, the shampoo will go on but will not be washed off…I believe that we are at the moderate stage at present.” | nfvPPA |
| **Theme 2: Illness progression/ trajectory** | |  |
| Fluctuations in decline | “One thing that has been very noticeable with my wife’s condition is that changes happen very quickly sometimes from one day to the next. It is not a gradual decline. However, once the drop off has happened I have on occasions been able to get my wife to recover some of the loss. It appears to me as though the brain is trying to find other ways of trying to get around the problem that it has been faced with.” | nfvPPA |
|  | “In our case there have been quite long periods of stability, then a sudden worsening. Sometimes this seems to have been brought about by a change to routine or an outside event, a domestic problem (something breaking down), staying away from home even for a short period. The condition then settles down again but doesn’t return to what it was before.” | lvPPA |
| Speed of progression | “The progression seemed faster than for other people we came across with this illness. For us it was 3 years from diagnosis to death.” | nfvPPA |
|  | “It definitely seems like the first couple years are gradual – and it is hard to notice changes until you look back after a year and realise they can’t do that anymore, or have been struggling – like driving. Infections/ delirium can massively exacerbate the illness and speed it up.” | lvPPA |
| **Theme 3: Experience of doing the research** | |  |
| Difficulties answering questions on behalf of the plwPPA | “These points are from pure observation since my wife has not been able to speak since the moderate stage of the condition and when she could speak she would never accept there was anything wrong with her.” | nfvPPA |
|  | “Just to say that I did find it quite difficult to recall the detail as my wife’s PPA has progressed and as a consequence I needed to make a sometimes not too educated guess about the period of time that my wife has spent on each stage.” | svPPA |
| Difficulties with the way the survey was designed | “I found it difficult to complete this questionnaire, not necessarily for emotional reasons, more because of the requirement to allocate the ‘correct’ positioning of the various symptoms to a particular stage.” | nfvPPA |
|  | “I found it very difficult to answer many questions, as the descriptions given of the proposed stages rarely corresponded to the actual progression of my partner (or of other friends living with PPA) and seemed to be based on pre-existing assumptions about the course and symptoms of the disease. Has any thought been given to using free descriptive text? I could more easily write a descriptive, chronological narrative. I really question the methodology used here.” | lvPPA |
| **Theme 4: Utility of the stages** | |  |
| Perceived strengths of the stages | “This is SO USEFUL & helpful for others as they embark on this challenging journey. Even if everyone experiences different symptoms at different rates, it’s so helpful as a carer to read and realise/ understand that these are ‘normal/expected’ with these diagnoses…you are not alone, weird, or must feel ashamed. Thank you – hugely appreciated from a carer’s point of view – so much if not all of it lands squarely on our shoulders!” | svPPA |
|  | “The stages so far are what I have experienced, no one ever told me the likely stages, it is left to carers to search for stages. This makes it very difficult to cope with and to prepare for. Describing the stages is a good idea and will be helpful for many carers.” | nfvPPA |
| Perceived limitations of the stages | “Regarding staging, the difficulty is like trying to decide where the boundaries lie between yellow, orange and red in the rainbow - making sharp boundaries between items on a continuous 'spectrum' can only be approximate. But I understand the need to try!” | lvPPA |
|  | “Stages make it sound very organised and predictable but, unlike Alzheimer’s perhaps, the progression of PPA isn’t like that. You can’t predict what will happen next or when it will happen, only that it is likely to happen some when. It sounds very harsh but this is a cruel disease.” | nfvPPA |
| **Theme 5: Suggestions for further development/dissemination** | |  |
| Incorporating care milestones/ appropriate therapies into the stages | “To then match the stages to actions which may be required by the patient and their carers, e.g. to sort out legal matters, Wills, Powers of Attorney, Guardianship, Advanced Care Directives etc at the earliest possible Stages. When to stop driving, when assistance with self care, assistance with activities of daily living, 24 hour care etc may be required (rough guides accounting for individual variations, valuable to carers nonetheless…)” | nfvPPA |
|  | “Reference to types of therapies that may be helpful at later stages – input from neuro physios and neuro occupational therapists so that appropriate physical and other sensory therapies can be used when other activities become too difficult or do not maintain interest. Thank you for doing this.” | lvPPA |
| Aligning stages with intact abilities | “I wonder if it might be useful to find out what people can do at different stages for the different dementias rather than what is declining…not so interesting for you medically speaking but very useful sharing information for carers. Making things meaningful and purposeful for both carer and caree is so important and may make life more meaningful/ rewarding/ satisfying.” | svPPA |
|  | “I think this is great but maybe would be also useful to add what the person IS still able to do as well as CAN’T.” | lvPPA |
| Acknowledging individual differences | “Maybe something indicating the percentage likelihood of having each particular symptom would be useful as mum hasn’t got every symptom although quite a few.” | svPPA |
|  | “I dare say everyone’s progress through the illness is different, and I’m sure you will make this clear in your leaflet.” | lvPPA |
| Importance of how and when information is accessed | “I think the points made in the introduction are very valid – a road map of symptoms presented at an early stage could well be overwhelming and distressing to contemplate. A partner may feel unequal to the task of managing these symptoms when they are described in behavioural terms. The person with the diagnosis may feel life would not be worth living with these symptoms”. | svPPA |
|  | “I have thought hard about whether it would be useful to inform people that they do not have to read all the stages but limit themselves to the stage they think the person is at. I think both of us would have been quite devastated and found it hard not to become depressed if we had had all the stages mapped out as will now be available. We have found it easier to deal with things as they came up. It certainly would have been very helpful for me, especially as the carer, to know what the next stage would entail but I would not have wanted to go further. I understand that this is a personal view and not everyone will think the same. Some may want to know the whole of what is to come, but that should be a conscious decision not one done accidentally.” | svPPA |

The Table is the fully expanded version of Table 3 in the main manuscript, and presents Themes and Subthemes identified in the Qualitative Framework Analysis, with illustrative caregiver quotations representing each Subtheme. lvPPA, logopenic variant primary progressive aphasia; nfvPPA, nonfluent/agrammatic variant primary progressive aphasia; plwPPA, person living with primary progressive aphasia; svPPA, semantic variant primary progressive aphasia

**Supplementary Table S5. Spanish translation of Table 1.**

We had originally included a Spanish translation of Table 1 here. MedRxiv did not allow us to upload this, advising the following: “According to our policies, any information presented in a language other than English should be removed. We understand that such materials can still be an important part of your study. Therefore, if full translation is not possible, you may want to remove these materials and advise that readers contact the corresponding author to request access to this information.”

Please contact if you would like to see the Spanish translation of Table 1.
